# Microbiome Preterm Birth DREAM Challenge: Crowdsourcing Machine Learning Approaches to Advance Preterm Birth Research

**DOI:** 10.1101/2023.03.07.23286920

**Authors:** Jonathan L. Golob, Tomiko T. Oskotsky, Alice S. Tang, Alennie Roldan, Verena Chung, Connie W.Y. Ha, Ronald J. Wong, Kaitlin J. Flynn, Antonio Parraga-Leo, Camilla Wibrand, Samuel S. Minot, Gaia Andreoletti, Idit Kosti, Julie Bletz, Amber Nelson, Jifan Gao, Zhoujingpeng Wei, Guanhua Chen, Zheng-Zheng Tang, Pierfrancesco Novielli, Donato Romano, Ester Pantaleo, Nicola Amoroso, Alfonso Monaco, Mirco Vacca, Maria De Angelis, Roberto Bellotti, Sabina Tangaro, Abigail Kuntzleman, Isaac Bigcraft, Stephen Techtmann, Daehun Bae, Eunyoung Kim, Jongbum Jeon, Soobok Joe, The Preterm Birth DREAM Community, Kevin R. Theis, Sherrianne Ng, Yun S. Lee Li, Patricia Diaz-Gimeno, Phillip R. Bennett, David A. MacIntyre, Gustavo Stolovitzky, Susan V. Lynch, Jake Albrecht, Nardhy Gomez-Lopez, Roberto Romero, David K. Stevenson, Nima Aghaeepour, Adi L. Tarca, James C. Costello, Marina Sirota

## Abstract

Globally, every year about 11% of infants are born preterm, defined as a birth prior to 37 weeks of gestation, with significant and lingering health consequences. Multiple studies have related the vaginal microbiome to preterm birth. We present a crowdsourcing approach to predict: (a) preterm or (b) early preterm birth from 9 publicly available vaginal microbiome studies representing 3,578 samples from 1,268 pregnant individuals, aggregated from raw sequences via an open-source tool, MaLiAmPi. We validated the crowdsourced models on novel datasets representing 331 samples from 148 pregnant individuals. From 318 DREAM challenge participants we received 148 and 121 submissions for our two separate prediction sub-challenges with top-ranking submissions achieving bootstrapped AUROC scores of 0.69 and 0.87, respectively. Alpha diversity, VALENCIA community state types, and composition (via phylotype relative abundance) were important features in the top performing models, most of which were tree based methods. This work serves as the foundation for subsequent efforts to translate predictive tests into clinical practice, and to better understand and prevent preterm birth.

## Introduction

Preterm birth (PTB) is the leading cause of infant morbidity and mortality worldwide. Globally, every year approximately 11% of infants every year are born preterm, defined as birth prior to 37 weeks of gestation, totaling nearly 15 million births^1^. In addition to the emotional and financial toll on families, preterm births result in higher rates of neonatal death, nearly 1 million deaths each year, and long-term health consequences for some children^2^. Infants born preterm are at risk for a variety of adverse outcomes, such as respiratory illnesses, cerebral palsy, infections, and blindness, with infants born early preterm (i.e., before 32 weeks) at increased risk of these conditions^3^. Thus, the ability to accurately identify women at risk for PTB is a first step in the development and implementation of treatment and prevention strategies. Currently, available treatments for pregnant women at risk of preterm delivery include corticosteroids for fetal maturation and magnesium sulfate provided prior to 32 weeks to prevent cerebral palsy^2^. Progesterone supplementation may also be administered as early as the second trimester to reduce the risk of PTB^4^.

There are several known factors associated with PTB, including history of PTB, a short cervix, extremes of maternal age and body mass index (BMI), low socio-economic status, smoking, and genetic polymorphisms^5–11^. Nevertheless, there are currently no clinical tools that enable the early and reliable assessment of the risk of preterm birth for an individual^12, 13^. Machine learning (ML) modeling has demonstrated potential to aid in the determination of individuals at risk of conditions and diseases across medical domains^14–16^. By applying ML methods to large amounts of heterogeneous data, patterns in data can be discerned that would be otherwise difficult for humans to distinguish. Moreover, deducing which features contribute most to the predictive performance of an ML model allows for the identification of biomarkers that can be important for a condition or disease. There are a variety of ML algorithms that can be used individually, or combined into an ensemble approach to improve prediction performance. After ML modeling has been applied to and optimized on a training dataset, then the model is ideally tested on an independent dataset to assess how well the model is able to generalize to data it has never seen before^17^. The validation on independent data is a critical step to guard against overfitting and hence optimistically biased accuracy estimates. In the past several decades, applications of machine learning approaches to various types of clinical, molecular, and other data have been explored to predict complications of pregnancy including preterm birth^18–23^. The results of these works to date demonstrate that the prediction of PTB from varied data types including metabolites in amniotic fluid and maternal blood and urine, ultrasound images, and electronic health records, appears to be feasible to a certain extent. In 2019, a DREAM (Dialogue for Reverse Engineering Assessments and Methods) Challenge was organized to harness the power of crowdsourcing and engage the computational biology community to develop and apply machine learning models to maternal blood multi-omics data for the determination of gestational age at time of blood draw and prediction of spontaneous PTB^24^. Tarca et al.^24^ demonstrated that models based on the maternal blood transcriptome were able to significantly predict a subset of spontaneous PTBs (preterm prelabor rupture of the membranes) while spontaneous preterm labor and delivery was significantly predicted by the plasma proteome. For both outcomes, the sample closer to delivery was more informative than earlier samples.

Although the sources of some data to which ML algorithms can be applied are more difficult to obtain, such as blood and amniotic fluid which involve procedures that require technical expertise and puncturing through skin and other anatomical structures that may introduce infection or cause pain, vaginal microbiome samples can be collected relatively more easily by clinicians as well as by patients themselves^25^. There is some indication that the vaginal microbiome is associated with adverse pregnancy outcomes, specifically PTB.

Previous studies have shown that there are significant differences between the vaginal microbiome of patients who deliver at term and those who deliver prematurely. Vaginal microbiomes with increased diversity as well as communities where *Lactobacillus* is not dominant were more frequent in patients with PTB^26–28^. Therefore, the vaginal microbiome is a tempting source of data to use for predictive modeling of PTB. However, there are significant biological and technical challenges to using microbiome data for predictive modeling. Biologically, human-associated microbiomes (including the vaginal microbiome) are incredibly variable–with any two individuals typically sharing less than half of microbes^29^. Thus, microbiome data, particularly compositional microbiome data, is both highly dimensional (typically 10 to 100 times more features than biological replicates being observed) and sparse (most features are observed in few biological replicates). These microbiome data attributes contribute to a substantial risk of model overfitting. Metaanalysis as well as rigorous evaluation of models on independent validation data is a robust approach to contend with these biological challenges with microbiome data. However there are significant technical challenges in aggregating and combining microbiome data across studies, therefore there have been few studies taking on this task^30–32^. In previous work, we have shown that by aggregating microbiome data across several studies we can gain significant statistical power to show that higher diversity is associated with PTB especially in the first trimester of pregnancy and to identify several novel microbial associations^33^. While ML approaches have been applied to the vaginal microbiome, most have involved a single dataset with limited sample size^34–36^. One recent work explored the application of ML to 12 vaginal microbiome datasets to predict PTB; however, while they leveraged public data extensively to ensure their findings were robust across studies, their work did not include an independent validation dataset^30^. Moreover, their work involved a single approach - a random forest ML model - with predictive accuracy for PTB ranging from 0.28 to 0.79. We hypothesized that applying advanced computational and machine learning techniques to aggregated microbiome data across many diverse studies could be used successfully for identification of women at risk of delivering preterm, including against independent validation data unavailable to the models in the training phase.

Building on the groundwork laid by the 2019 Preterm Birth Transcriptome Prediction DREAM Challenge^24^, we designed a new Challenge aimed at leveraging longitudinal microbiome data and crowdsourcing for prediction of (i) preterm or (ii) early PTB. DREAM Challenges define the prediction task, supply the necessary data, and provide the infrastructure to evaluate models designed by any participating teams; they do so in an unbiased manner using a gold-standard, undisclosed validation dataset. The Challenges are international, open science efforts to identify the best predictive models. Here, we provide the results from the Preterm Birth Microbiome Prediction Challenge, along with top models, and insights gained from this initiative. The dockerized code for all predictive pipelines are made available along with data used in the challenge at: http://www.synapse.org/preterm_birth_microbiome. This work can serve as the foundation for subsequent endeavors to better understand the mechanisms underlying PTB and early PTB, to translate into clinical practice predictive tests to help identify women at risk of delivering preterm, and to discover interventions for prevention of PTB. Likewise, we believe this is a robust scientific approach suitable for predictive modeling of other conditions based on microbiome data.

## Results

### Overview

The overall timeline of the Microbiome PTB DREAM challenge is shown in Figure 1. Major milestones included developing and harmonizing the training data, opening of the challenge to participants, post-hoc integration and harmonization of the validation data, assessment of models, and finally evaluation of the approaches and results. We leverage data across 9 studies including over 3,500 samples and utilized crowdsourcing to identify best predictive strategies and models for prediction of PTB. The endpoints of the challenge included PTB (delivery before 37 weeks of gestation) and early PTB (delivery before 32 weeks of gestation).

**Figure 1:**
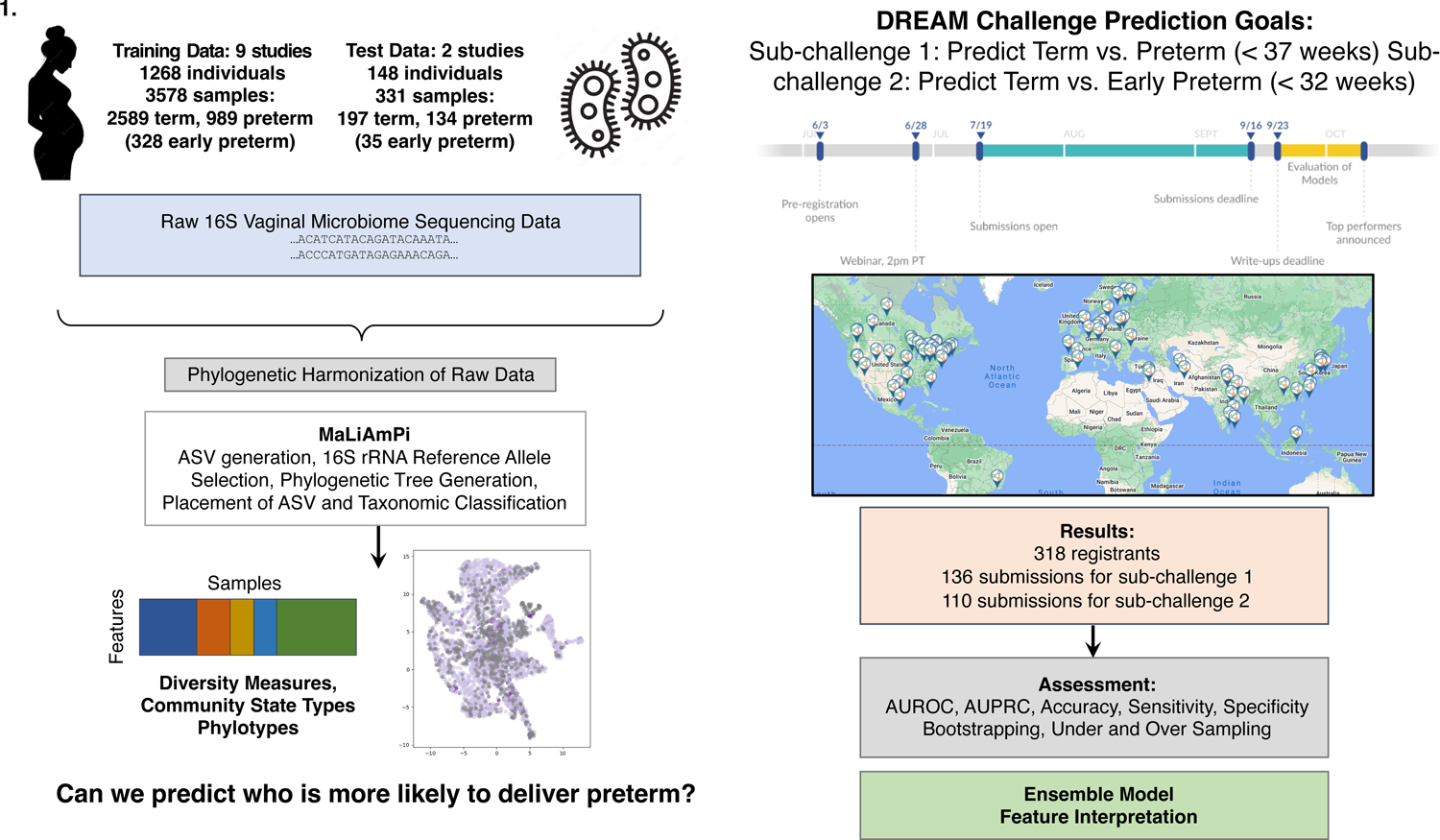
Study Design and Challenge Overview

### Data Aggregation and Processing

The training dataset was constructed by aggregating and processing vaginal microbiome data from the public domain leveraging resources including dbGAP^37^ as well as MOD Database for Preterm Birth Research^38^. The final dataset included data from nine studies, representing 3,578 samples from 1,268 individuals. Of these patients, 851 delivered at term and 417 preterm (before 37 weeks of gestation) including 170 whose deliveries were early preterm (before 32 weeks of gestation). Details of the nine studies that were included in the training set are shown in Table 1. Supplementary Figure 1 illustrates the sampling strategies for each of the datasets, showing that some studies (like I and J) collected samples only once during gestation, while in most other studies samples were collected multiple times during gestation from the same individual. As shown in Table 1, while all of these studies focus on profiling the 16S rRNA gene, primers targeting different variable regions of the 16S rRNA gene, PCR conditions, and sequencers all varied. The combination of microbiome data from different studies, particularly those using different underlying techniques, is a challenging task which has hindered prior efforts for meta-analysis of microbiome data. Likewise, integration of newly sequenced microbiome data *ad hoc* into an existing set of features is another barrier to the practical use of microbiome-trained predictive models. This was evident when we generated our first ordination of the training and validation data based on raw sequence reads, all preprocessed with DADA2 into amplicon sequence variants (ASVs), where specimens clustered more by the underlying technique (Figure 2a), such as primer selection, variable regions amplified, and sequencing platforms used. Thus, we first focused on harmonizing the microbiome data from the nine studies that comprised our training set into a common set of features that were not reliant upon taxonomy, but instead based on phylogenetic placement of the ASVs onto a common *de novo* maximum likelihood phylogenetic tree comprised of full-length 16S rRNA alleles. This approach is fully described and validated elsewhere, and was implemented as a Nextflow-based workflow called MaLiAmPi^39^. After processing with MaLiAmPi, we were able to overcome most of the technique-based noise and successfully harmonize the data into one cohesive feature set. As seen in Figure 2b, phylogenetic placement resulted in Shannon alpha diversity measures that were consistent across the majority of the studies after processing with MaLiAmPi, although study F did have higher diversity across the samples. The separation between samples by outcome–from term, preterm, and early preterm deliveries–is not clearly evident (Figure 3a and b). There are some distinct differences observed with respect to community state types (CSTs) and outcome (Figure 3c and Supplementary Figure 3). Leveraging different types of microbial features including phylotype relative abundance, diversity measures as well as CST membership provide a unique opportunity to apply ML techniques to these data for PTB prediction. Additional dimensionality reduction plots demonstrating the successful integration of the data, colored by trimester of collection and demographic features, are presented in Supplementary Figure 2.

**Figure 2.**
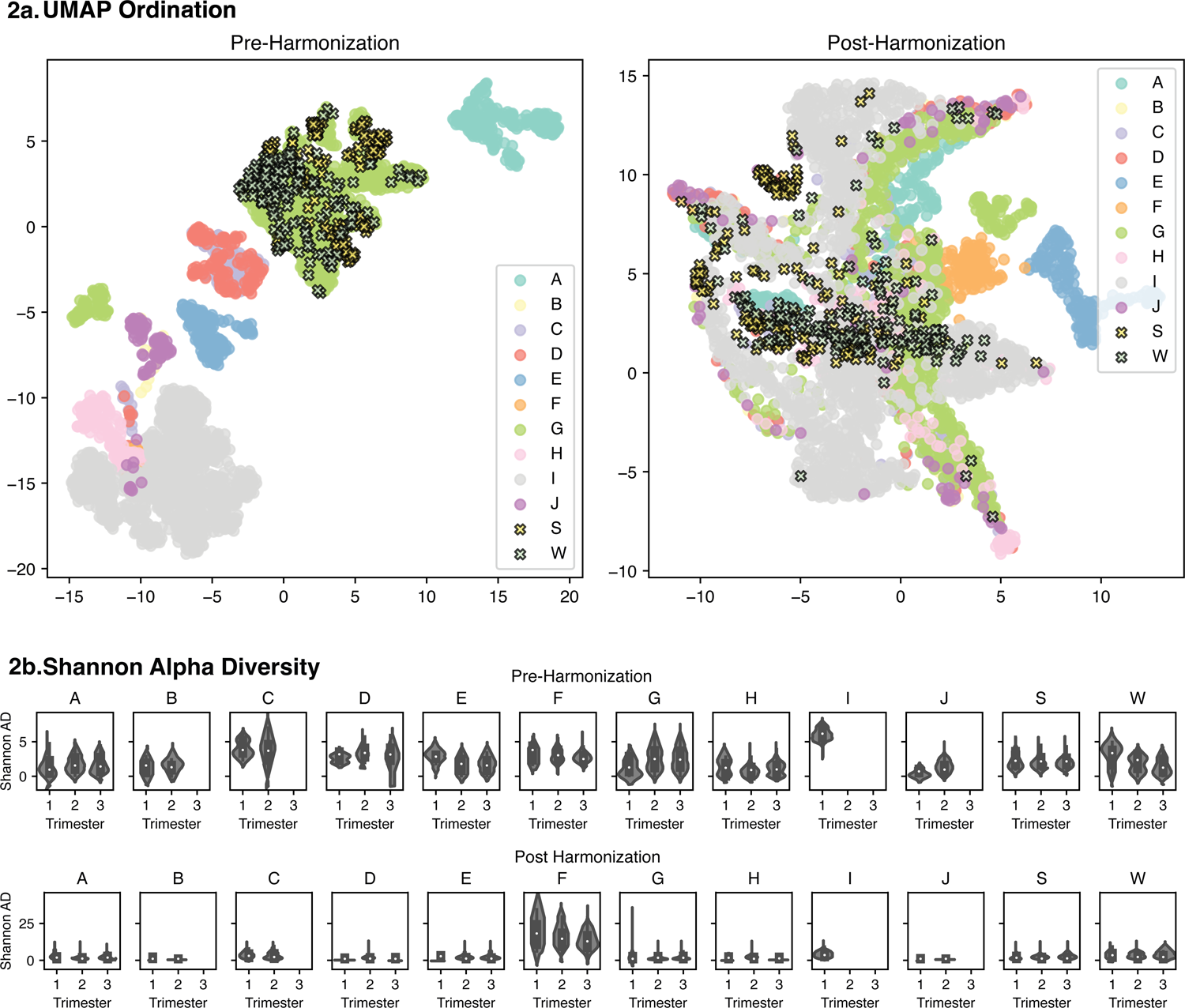
Data visualization of harmonization by Maliampi of microbiome data across studies. a) Uniform Manifold approximation and projection (UMAP) ordination plots of the aggregated data before (left) and after (right) harmonization where each dot represents one vaginal microbiome sample colored by study. b) Violin plots of Shannon alpha diversity by trimester before (top) and after (bottom) harmonization stratified by study

**Figure 3.**
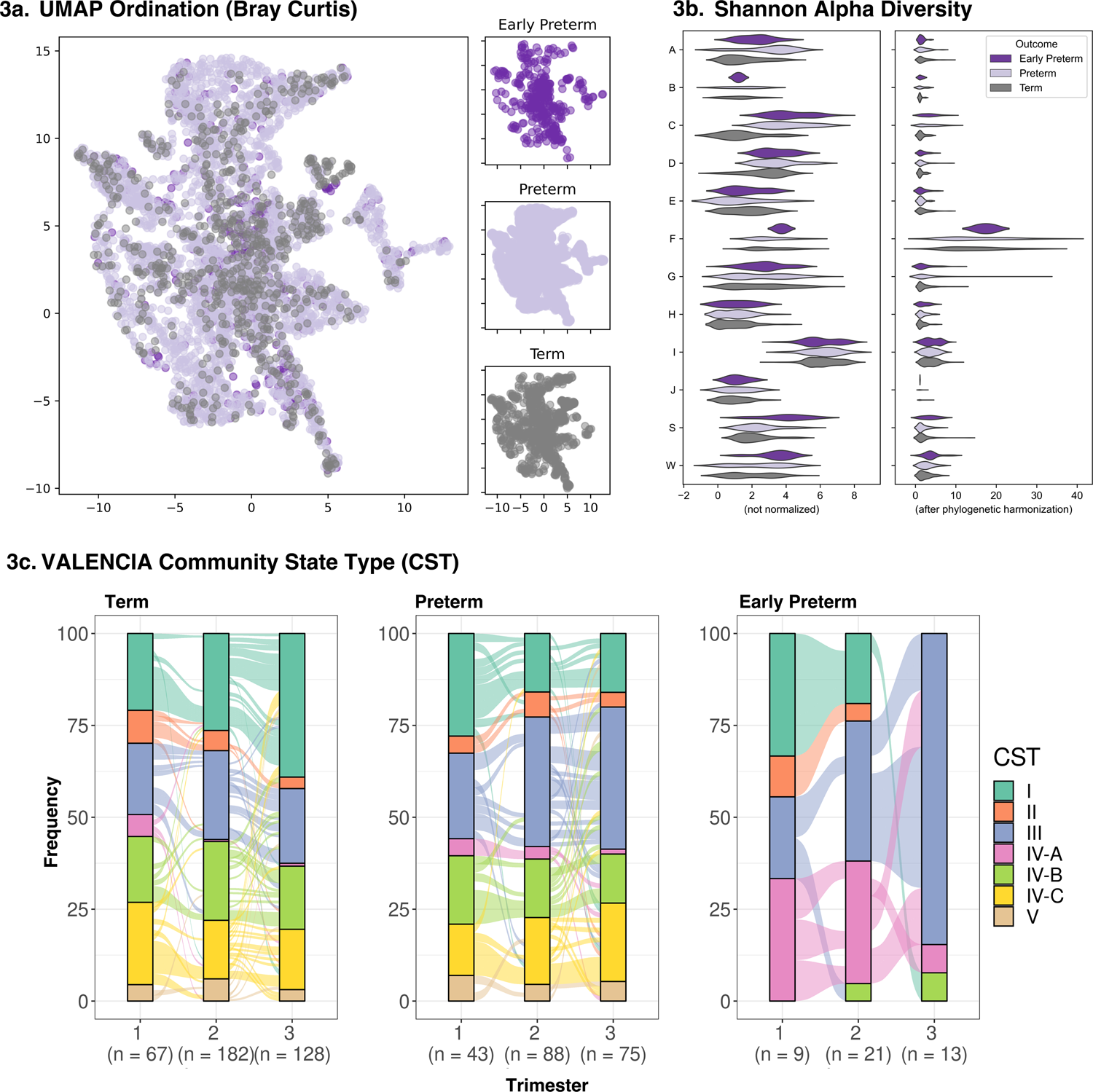
Data visualization of microbiome features by outcome. a) Uniform Manifold approximation and projection (UMAP) ordination plots of the vaginal microbiome colored by outcome, b) Violin plot of diversity before (left) and after (right) harmonization stratified and colored by outcome and c) Alluvial plot of community state type (CST) frequencies across time stratified by birth outcome

**Table 1:** Summary of participants, samples, and V region sequences of training (AJ) and validation (W and S) datasets

| Study ID | Study Accession ID | Center | Title (Authors, year) | # of Participants | # of Term \| PTB \| Early PTB Participants | # of Samples | # of Term\|PTB \| Early PTB Samples | V Region Sequences | Instrument |
| --- | --- | --- | --- | --- | --- | --- | --- | --- | --- |
| A | SDY465 | Stanford University | Temporal and spatial variation of the human microbiota during pregnancy (DiGiulio et al., 2015) | 39 | 32\|7\|3 | 231 | 180\|51\|21 | V3 - V5 | 454 GS FLX Titanium |
| B & J | PRJEB11895 & PRJEB12577 | Imperial College London | The interaction between vaginal microbiota, cervical length, and vaginal progesterone treatment for preterm birth risk (Kindinger et al., 2017) | 116 | 91\|25\|9 | 116 | 91\|25\|9 | V1 - V3 | Illumina MiSeq |
| C | PRJEB21325 | Imperial College London | Vaginal dysbiosis increases risk of preterm fetal membrane rupture, neonatal sepsis and is exacerbated by erythromycin (Brown et al., 2018) | 110 | 18\|92\|49 | 144 | 20\|124\|67 | V1 - V2 | Illumina MiSeq |
| D | PRJEB30642 | Imperial College London | Establishment of vaginal microbiota composition in early pregnancy and its association with subsequent preterm prelabor rupture of the fetal membranes (Brown et al., 2019) | 70 | 15\|55\|21 | 134 | 26\|108\|38 | V1 - V2 | Illumina MiSeq |
| E | PRJNA242473 | University of Maryland | The vaginal microbiota of pregnant women who subsequently have spontaneous preterm labor and delivery and those with a normal delivery at term (Romero et al., 2014) | 73 | 57\|16\|10 | 168 | 137\|31\|19 | V1 - V3 | 454 GS FLX Titanium |
| F | PRJNA294119 | Washington University | Early pregnancy vaginal microbiome trends and preterm birth (Stout et al., 2017) | 74 | 51\|23\|2 | 145 | 99\|46\|2 | V1 - V3 | 454 GS FLX Titanium |
| G | PRJNA393472 | Stanford University | Replication and Refinement of a Vaginal Microbial Signature of Preterm Birth (Callahan et al, 2017) | 134 | 85\|49\|20 | 957 | 670\|287\|71 | V4 | Illumina HiSeq 2500 |
| H | PRJNA430482 | Virginia Commonwealth | The vaginal microbiome and preterm birth (Fettweis et al., | 114 | 70\|44\|11 | 216 | 137\|79\|19 | V1–V3 | Illumina HiSeq 4000 |
| I | PRJNA504518<br>(phs001739.v1.p1.) | University of Pennsylvania | Cervicovaginal microbiota and local immune response modulate the risk of spontaneous preterm delivery (Elovitz et al., 2019) | 538 | 432\|106\|45 | 1467 | 1229\|238\|82 | V3 - V4 | Illumina HiSeq 2500 |
| S | Not applicable | Stanford University | Not applicable | 88 | 48\|40\|8 | 172 | 95\|77\|18 | V4 | Illumina NextSeq 550 |
| W | Not applicable | Wayne State University | *Working title: The Vaginal Microbiota in Early Pregnancy Identifies a Subset of Women at Risk for Early Preterm Prelabor Rupture of Membranes and Preterm Birth* | 60 | 40\|20\|5 | 159 | 102\|57\|17 | V4 | Illumina MiSeq |
| Total | Not applicable | Not applicable | Not applicable | 1416 | 939\|477\|183 | 3909 | 2786\|1123\|363 | V1, V2, V3, V4, +/or V5 | 454 GS FLX Titanium, Illumina MiSeq, Illumina HiSeq 2500, Illumina HiSeq 4000, c Illumina NextSeq 550 |

To build an independent test set for evaluating the models submitted by participants in this DREAM challenge, we combined an unpublished dataset from Wayne State University consisting of 159 samples across 60 individuals among whom 40 (66.7%) had term deliveries and 20 (33.3%) had preterm deliveries, including 5 (8.3%) who had early preterm deliveries. Most patients in this test set had three longitudinal samples. We also generated a second validation dataset that comprised 172 vaginal microbiome samples from 88 individuals, up to three samples (one sample per trimester) for each individual, with 48 individuals (54.5%) having term deliveries, and 40 individuals (45.5%) having preterm deliveries including 8 (9.1%) having early preterm deliveries. DNA extraction, V4 16S rRNA gene library preparation, and 16S rRNA gene sequencing (2×150 Paired-End sequencing on the Illumina NextSeq platform) of these samples was performed by the UCSF Benioff Center for Microbiome Medicine, with most samples yielding over 100,000 reads (see Methods for details). Supplementary Figure 1 represents the week of gestation for the sample collection times for each individual from the two test datasets. These validation datasets became available only after the training dataset was generated and distributed to teams. Thus, the resultant reads had to be integrated into the same feature set as in the training data *post-hoc*. Using MaLiAmPi, we were able to first generate the training data, preserving the features (e.g., phylotypes, alpha diversity, etc.) (Figure 2a, B) and further integrate the validation datasets. The generalizability of these features across studies, including new study data, has allowed us to apply the ML models to these independent validation sets, and enable the use of the model on data to be generated in the future.

### The DREAM Challenge Results

The Preterm Birth Microbiome Prediction DREAM Challenge launched on July 5, 2022 (Figure 1) and closed on September 16, 2022. There were two sub-challenges for this challenge: sub-challenge 1 - Prediction of PTB (before 37 weeks of gestation) and sub-challenge 2 - Prediction of *early* PTB (before 32 weeks of gestation). The validation dataset for this second sub-challenge included only data from samples collected no later than 28 weeks of gestation (to reduce trivial predictions based upon later-in-gestation specimens being available from a pregnancy). A baseline ‘organizers’ random-forest based model was developed with the training data to provide participants an example, inclusive of packing of the model within a docker container. Performance metrics that were used to evaluate the prediction models submitted by the teams include area under the receiver operator characteristic (AUROC) curve, area under the precision-recall (AUPR) curve, accuracy, sensitivity, specificity and Matthews Correlation Coefficient (MCC). All values were determined on bootstrapped validation data, with the mean bootstrapped value used to evaluate the model. The primary scoring metric was set at the onset to be AUROC, followed by AUPR to break ties.

There were 318 participants from all over the world with 136 and 110 submissions for sub-challenges 1 and 2, respectively. The prediction models with top-ranking submissions achieved mean bootstrapped AUROC scores of 0.688 and 0.868 respectively for the 2 sub-challenges (Figure 4, Supplementary Tables 1 and 2). Several techniques were carried out in order to ensure the robustness of the resulting rankings including test set label inversion, bootstrapping, oversampling, and undersampling (see Methods). The results are shown in Supplementary Figures 4 (sub-challenge 1) and 5 (sub-challenge 2).

**Figure 4.**
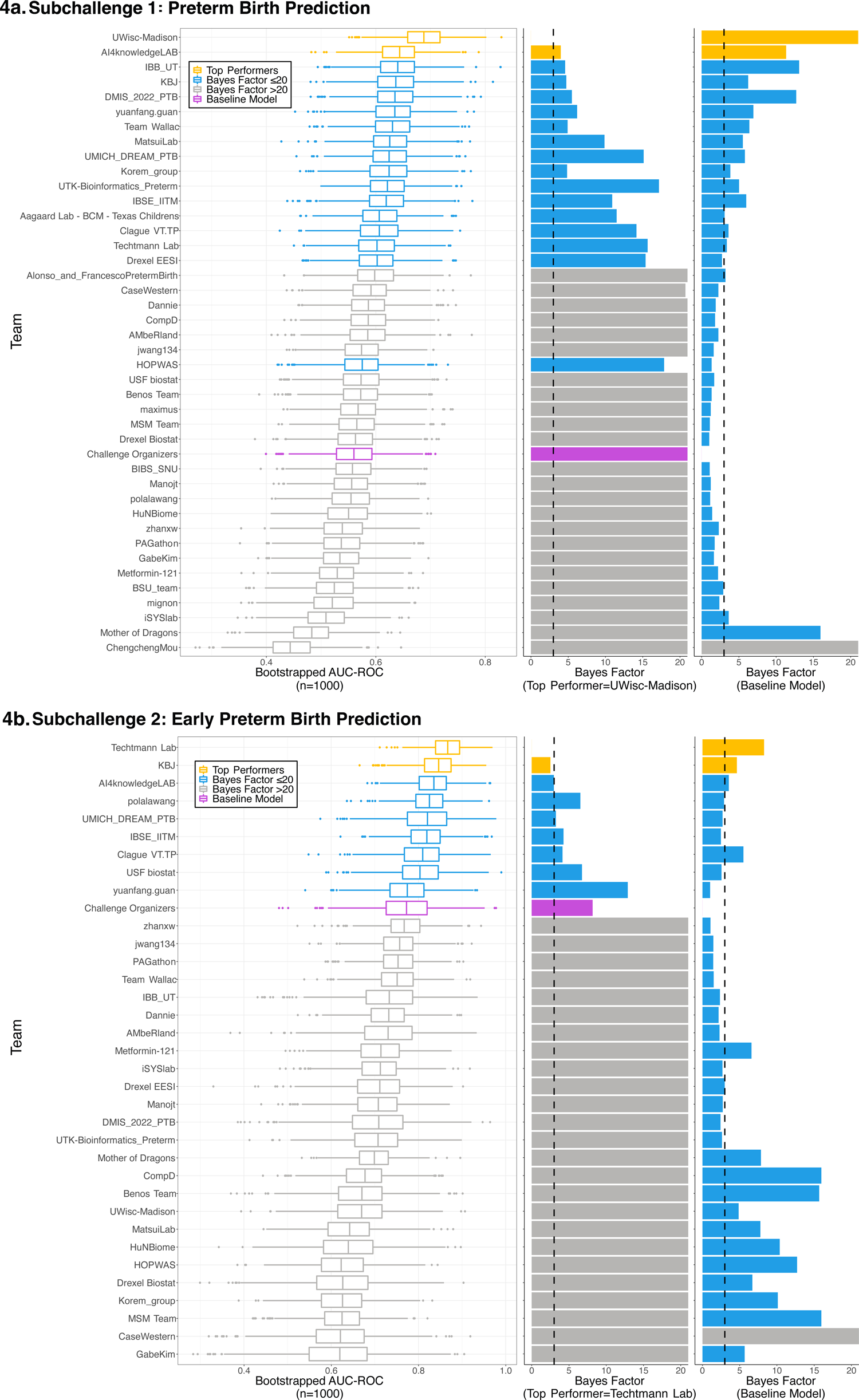
Challenge Results. Bootstrapped area under the receiver operator characteristics (AUROC) curves and Bayes factors for a) sub-challenge 1 and b) sub-challenge 2

**Table 2:** Summary of demographics of training (A-J) and validation (S and W) datasets

|  | Group | Total | Training (A - J) | Validation (S & W) |
| --- | --- | --- | --- | --- |
| **Individuals** | **n** | 1416 | 1268 | 148 |
| **Age Range, n (%)** | **Unknown** | 691 (48.8) | 691 (54.5) | 0 (0) |
|  | **Below 18** | 4 (0.3) | 4 (0.3) | 0 (0) |
|  | **18 to 28** | 304 (21.5) | 227 (17.9) | 77 (52.0) |
|  | **28 to 38** | 357 (25.2) | 293 (23.1) | 64 (43.2) |
|  | **Above 38** | 60 (4.2) | 53 (4.2) | 7 (4.7) |
| **Race, n (%)** | **Race: American Indian or Alaska Native** | 9 (0.6) | 6 (0.5) | 3 (2.0) |
|  | **Race: Asian** | 84 (5.9) | 81 (6.4) | 3 (2.0) |
|  | **Race: Black or African American** | 827 (58.4) | 759 (59.9) | 68 (45.9) |
|  | **Race: Native Hawaiian or Other Pacific Islander** | 7 (0.5) | 3 (0.2) | 4 (2.7) |
|  | **Race: White** | 422 (29.8) | 360 (28.4) | 62 (41.9) |
|  | **Race: Unknown** | 71 (5.0) | 63 (5) | 8 (5.4) |
| **Ethnicity, n (%)** | **Ethnicity: Hispanic or Latino** | 50 (3.5) | 8 (0.6) | 42 (28.4) |
|  | **Ethnicity: Unknown** | 1261 (89.1) | 1260 (99.4) | 1 (0.7) |
| **Delivery, n (%)** | **Term** | 939 (66.3) | 851 (67.1) | 88 (59.5) |
|  | **Preterm** | 477 (33.7) | 417 (32.9) | 60 (40.5) |
|  | **Early Preterm** | 183 (12.9) | 170 (13.4) | 13 (8.8) |

A few patterns emerged in the best-performing predictive models for sub-challenge 1 (Table 3) and subchallenge 2 (Table 4). Nearly all of the models used tree-based approaches (typically implemented as part of the python Scikit Learn^40^ package), such as random forest and relatives. A few models used regression approaches with inclusion of gestational age at sampling (with feature pruning and clustering), or neural networks. All of these modeling approaches are notable for their aggressive pruning or consolidation of features well-suited for handling both sparse and highly dimensional data. Therefore, avoiding overfitting the training data was a shared and likely essential attribute of the best-performing models.

**Table 3:**
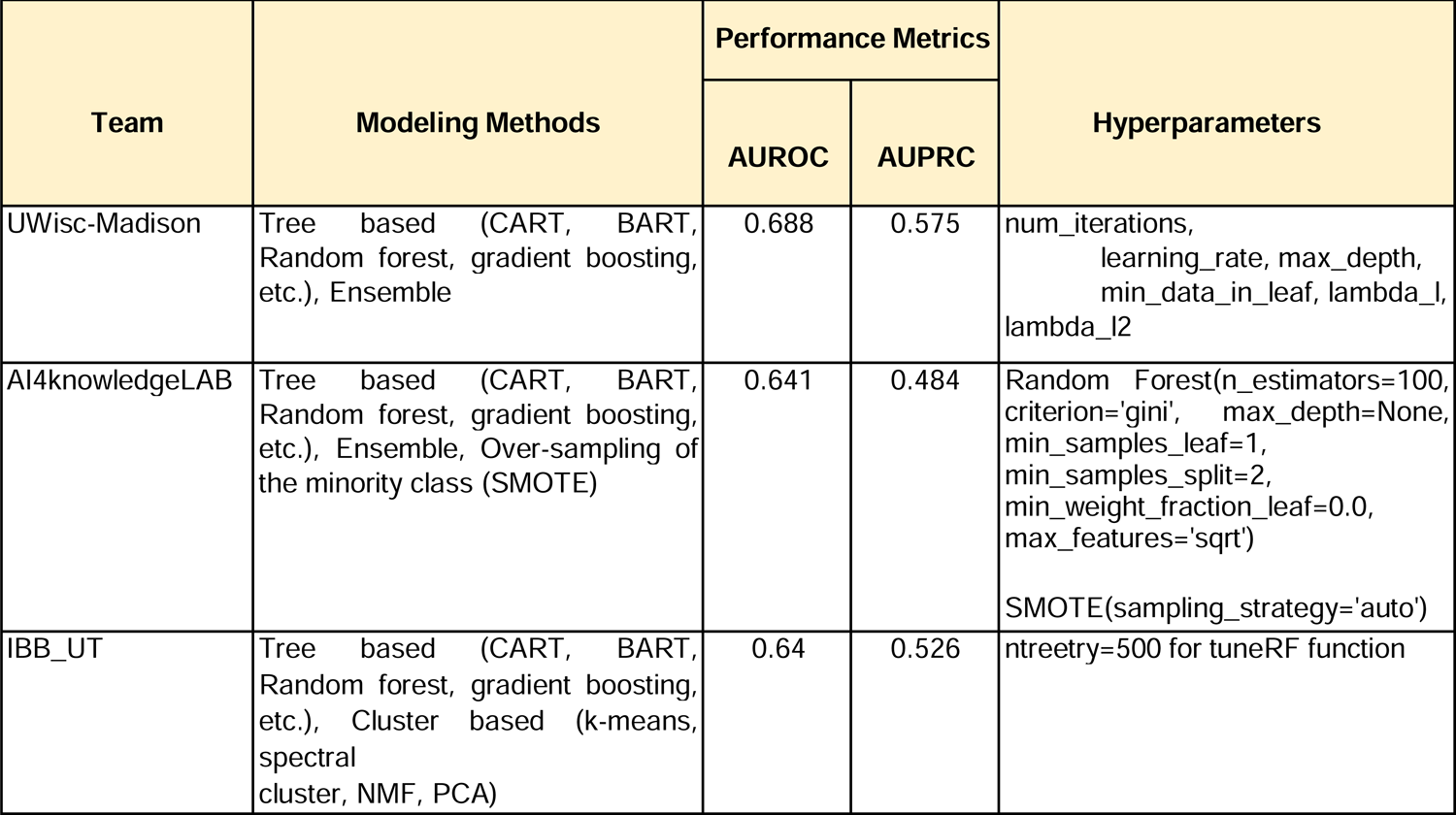
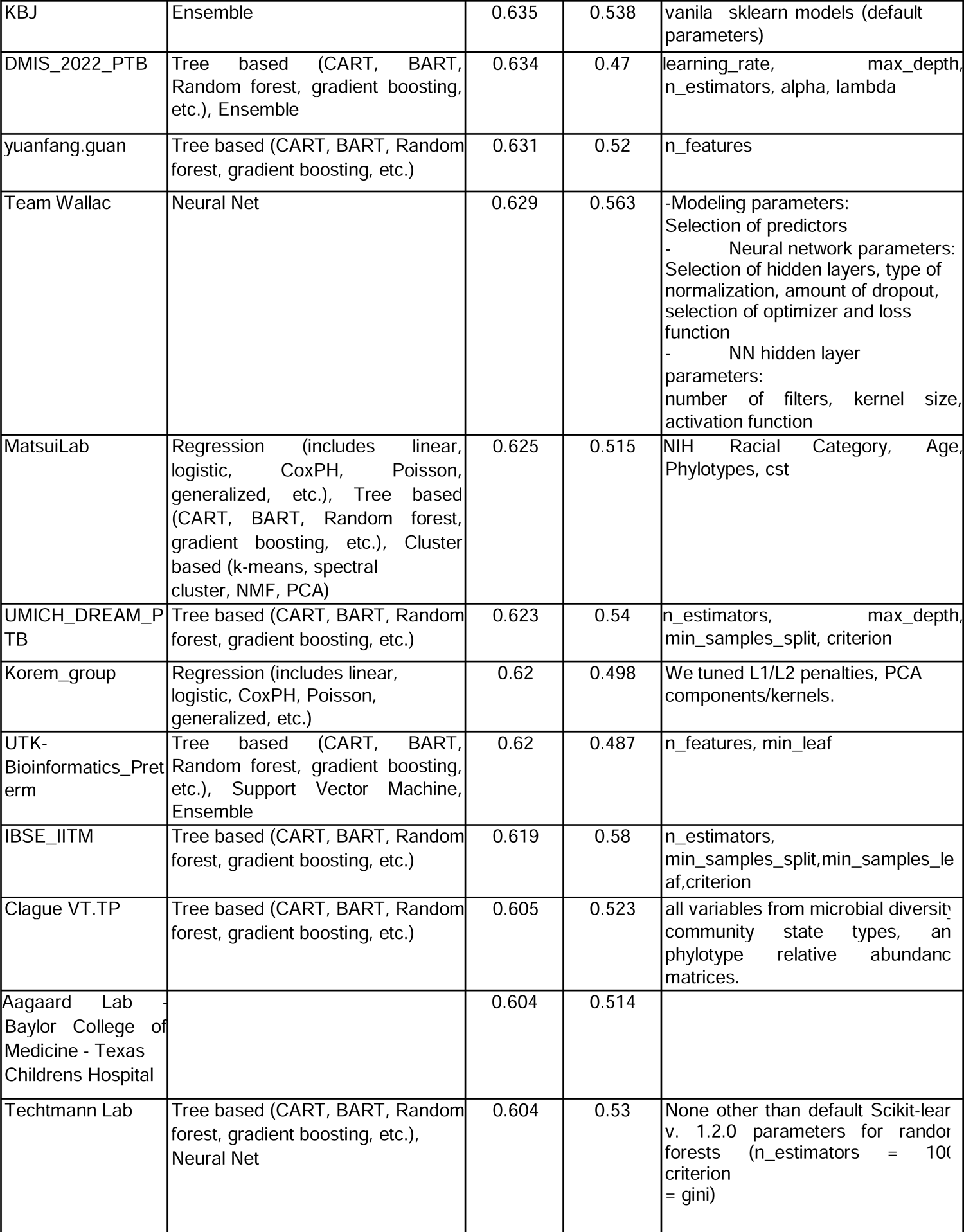
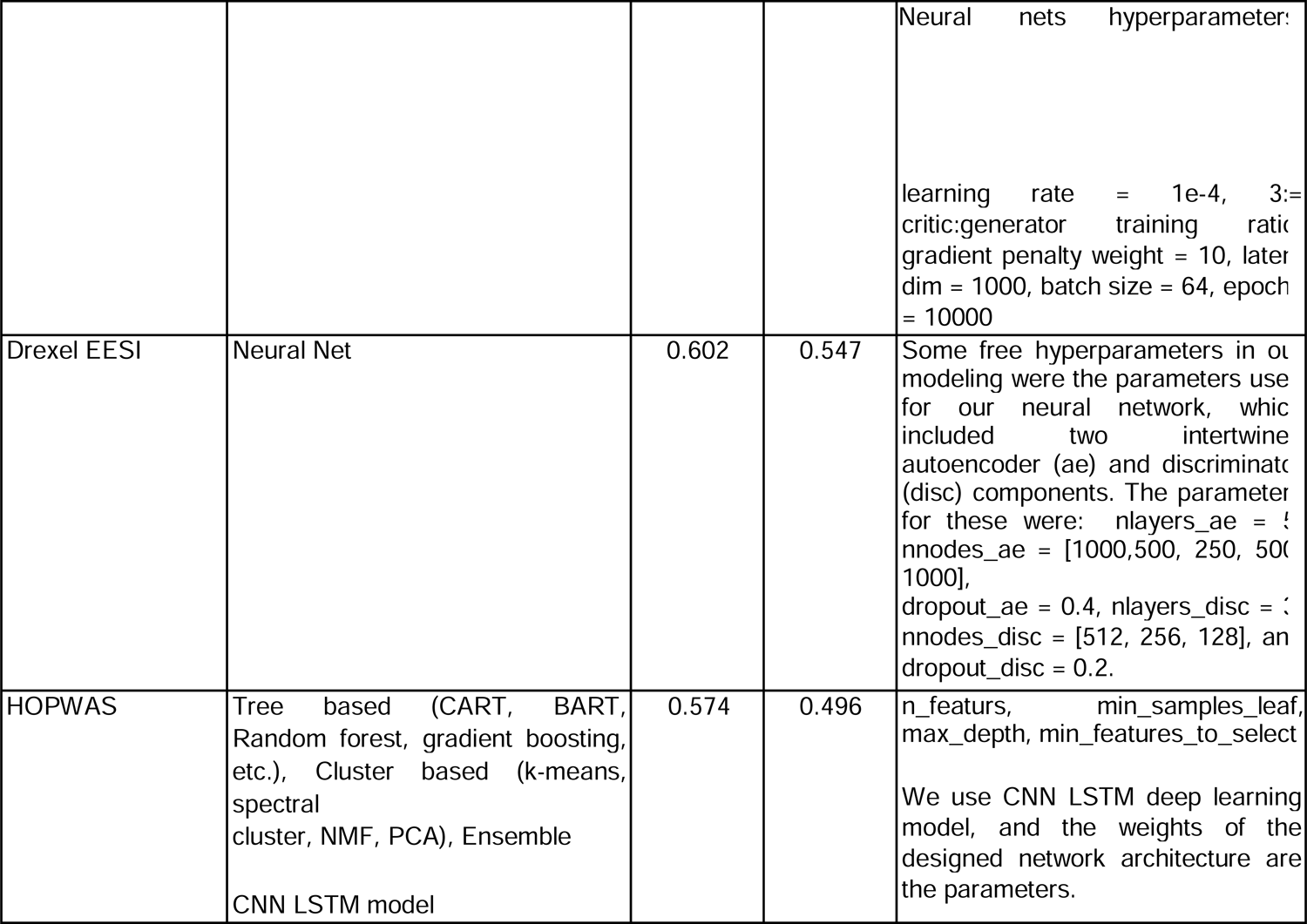
Summary for sub-challenge 1 of modeling methods, performance metrics, and hyperparameters for teams with Bayes factor < 20

**Table 4:**
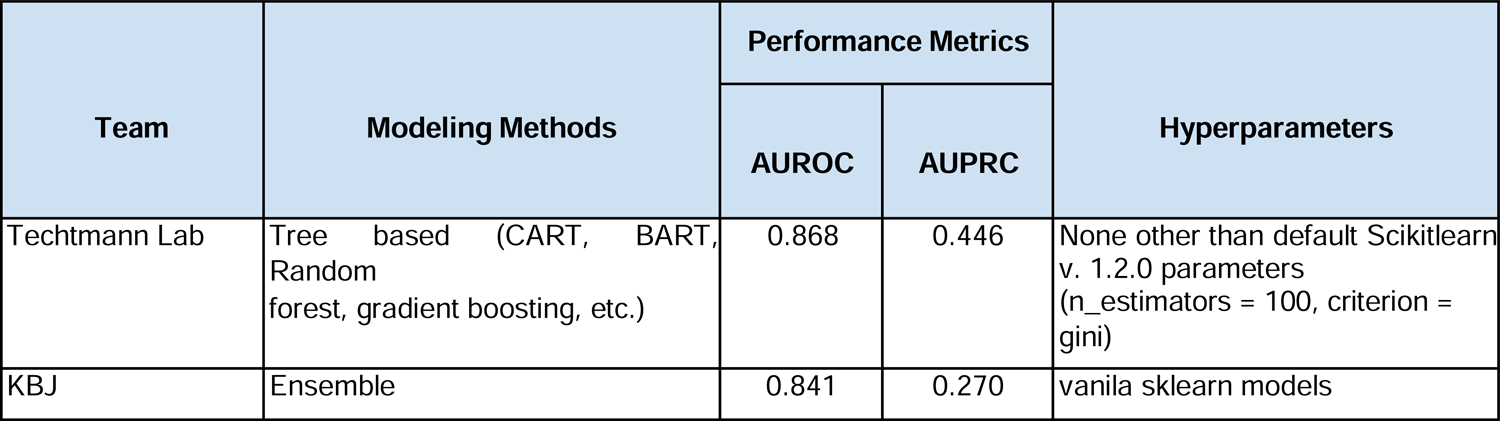
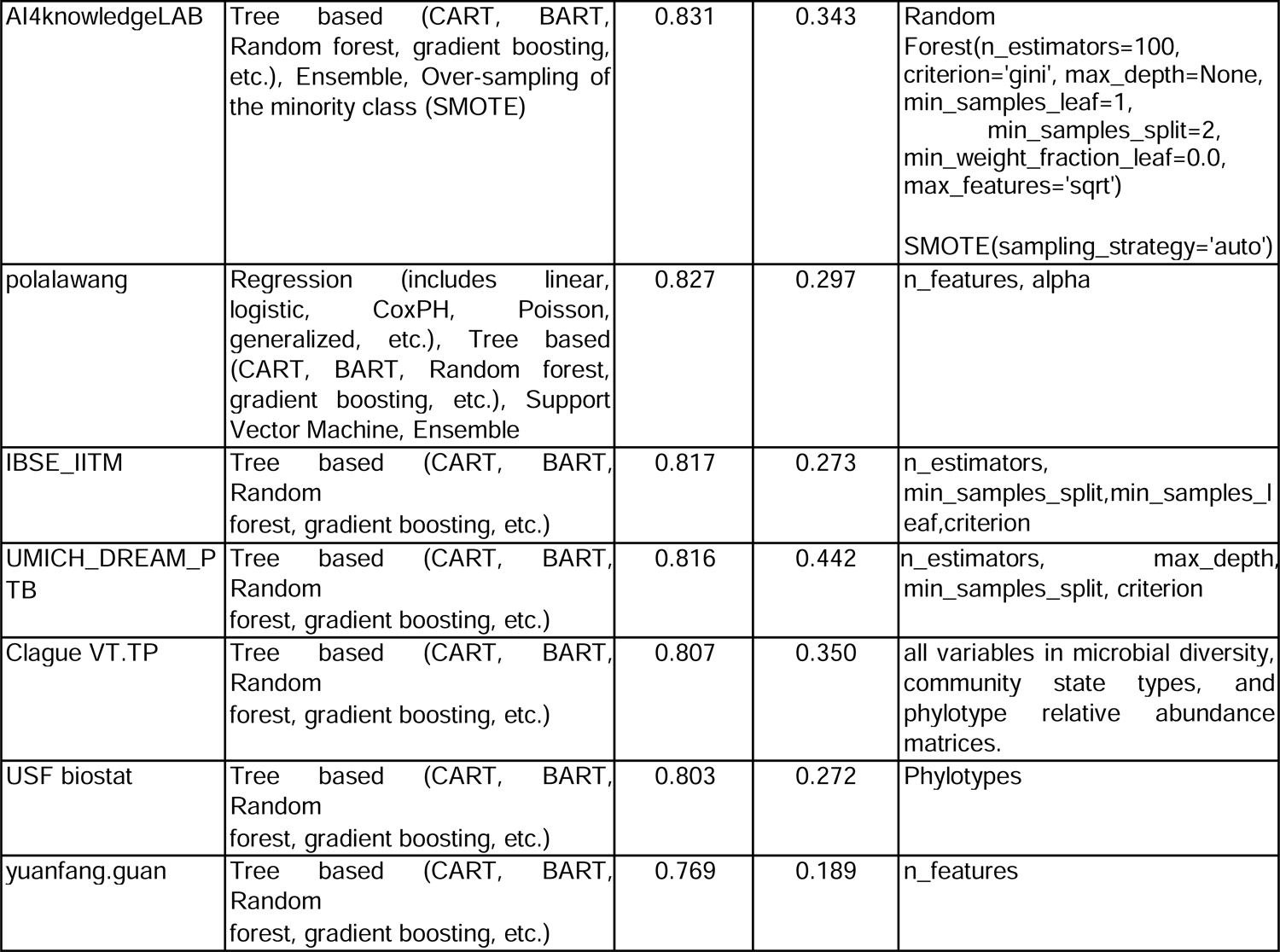
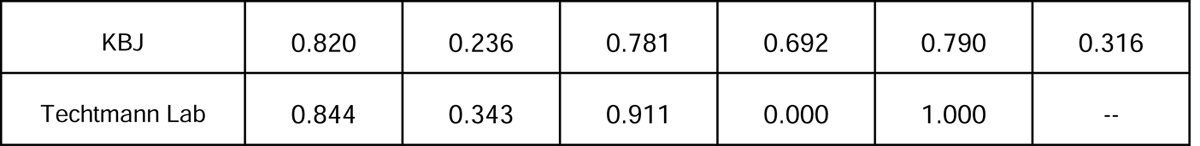
Summary for sub-challenge 2 of modeling methods, performance metrics, and hyperparameters for teams with Bayes factor < 20

### Predictive Features

Next we focused on identifying common features that the best performing models (as judged by mean bootstrapped AUROC, one model per team) relied upon to make their predictions. We used feature permutation (limited to models that could make a prediction in a tractable time) as a means of empirically identifying the feature tables and individual features that the models depended upon for their predictions. For both sub-challenges, the best performing models relied upon alpha diversity, VALENCIA community state types, and some form of composition (either phylotypes or taxonomy) (Figure 5). There was a preference for phylotypes over taxonomy for the very best performing models for both sub-challenges.

**Figure 5:**
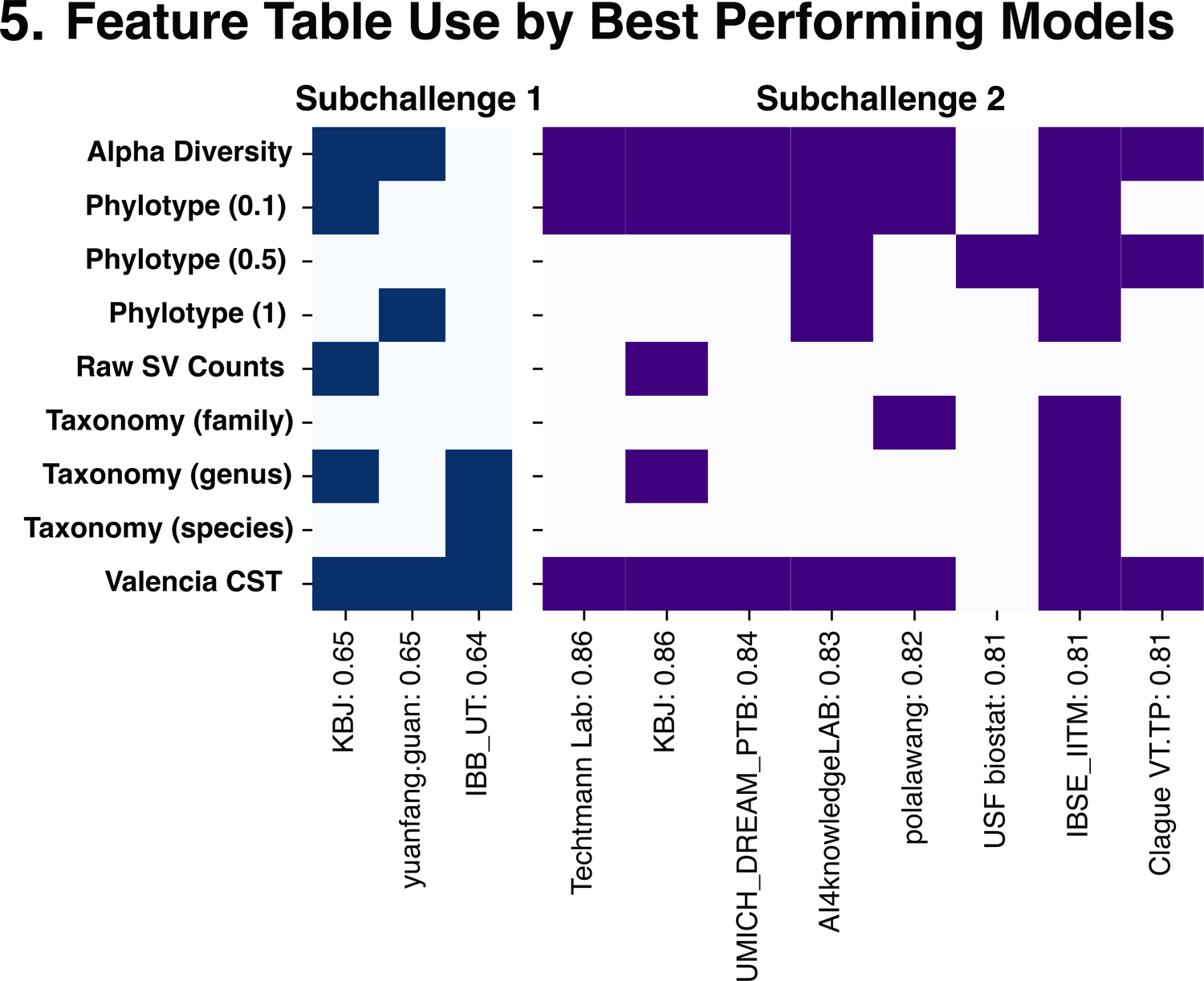
Feature Sets used by Top Performing Models. Feature tables used by the top performing models for sub-challenge 1 (left) and sub-challenge 2 (right)

We used feature permutation to first identify features used by the top-performing predictive models in subchallenge 1 (Figure 6a), and then proceeded to establish the univariate relationship with PTB stratified by trimester. A surprising number of phylotypes (at a phylogenetic distance of 0.1) were significantly associated with PTB in the second trimester (Figure 6a) when analyzed as present-absent and assessed with a Fisher’s exact test and contingency table after summarizing each pregnancy by trimester (to address repeated sampling in some of the underlying studies). As would be expected, *Lactobacillus* species generally were negatively associated with PTB. Curiously, one *Lactobacillus jensenii*-like phylotype is positively associated with PTB when present in the second trimester. Likewise, in the third trimester, two distinct *Lactobacillus* were more prevalent with PTB. Both are contrary to the broad notion that *Lactobacillus* are beneficial in preventing PTB. Alpha diversity metrics (Figure 6a) and VALENCIA community state types (Figure 6a) were largely insignificant when evaluated as univariates.

**Figure 6:**
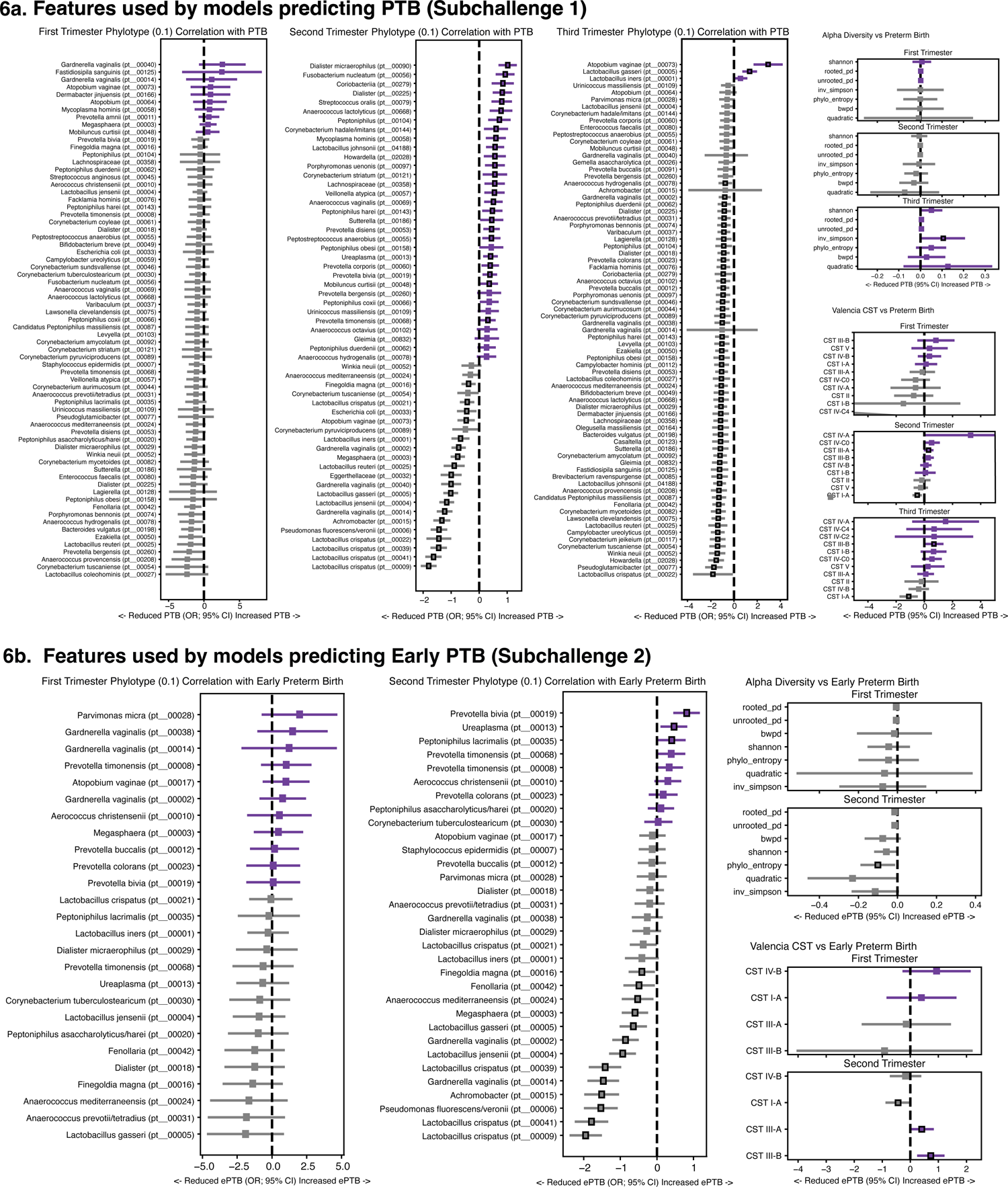
Features Across Best Performing Models. For models performing at threshold or above baseline, odds ratios (OR) with 95% confidence intervals (CI) reflecting correlation with PTB by trimester of specific phylotypes (0.1), diversity metrics, and community state types (CSTs) of features used extensively by top-performing models for a) sub-challenge 1 and b) sub-challenge 2

Much like with sub-challenge 1, we next used feature permutation to identify features used by multiple topperforming models in sub-challenge 2, predicting early PTB, followed by univariate correlation of these features with early PTB (Figure 6b). The better predictive performance of the sub-challenge 2 models (predicting early PTB) makes feature permutation more robust. In the second trimester (Figure 6b), phylotypes corresponding to multiple *Lactobacillus* strains were less prevalent in early PTB pregnancies. Curiously, one *Gardnerella vaginalis* strain was less prevalent in early PTB, contrary to this organism typically being thought of as a risk factor. For alpha diversity (Figure 6b), increased phylogenetic entropy in the second trimester was the most cleanly associated with early PTB. VALENCIA community state type IIIA or III-B in the second trimester were the most associated with early PTB (Figure 6b).

### Sub-challenge 1 - Top performing teams

#### Team UWisc-Madison

For predicting PTB, a LightGBM-based pipeline was built using an ensemble strategy tailored for vaginal microbiome data collected from multiple projects. The model was developed using specimens collected no later than 32 weeks of gestation and included five types of features: counts of taxa at different taxonomic levels, counts of phylotypes, microbiome community states, alpha diversity metrics, and metadata (age, collection week, and race). In particular, the counts of taxa at the family, genus, and species levels, the counts of phylotypes defined at phylogenetic distances of 0.5 and 1, and the alpha diversity metrics including Shannon index, Inverse Simpson Index, phylogenetic entropy, balance-weighted phylogenetic diversity, and rooted/unrooted/quadratic phylogenetic diversity were used. To obtain scale-invariant values, the centered log-ratio (CLR) transformation^41^ was applied to each type of the microbiome count data. Rare microbial features with less than 5 non zero counts in any of the studies of the training set were removed. The LightGBM model was chosen as the prediction model due to its well-known efficiency^42^. Each specimen was one training sample and each training sample had a total of 1,991 features. Five-fold cross-validation on the subject level was used to tune hyperparameters. Because Project G had a very different sequencing depth profile (the average sequencing depth of Project G is 185,010, whereas the value is below 50,000 for other projects), two prediction models were built: one was trained using specimens from all projects (Model 1) and one was trained only using specimens from Project G (Model 2). When making a prediction given a specimen, the ensembling weights of Model 1 and Model 2 were generated by a logistic regression model with sequencing depth and collection week as features. As one subject is likely to have multiple vaginal microbiome specimens, a customized weighting method was designed to aggregate predictions from multiple specimens on one subject. If a subject has multiple specimens, then the weight of each specimen equals the collection week of the specimen divided by the sum of the collection weeks of all specimens from the subject. In other words, the closer a sample was to delivery, the more impact it would make on the final prediction. The architecture of the pipeline is presented in Supplementary Figure 6. This pipeline achieved an AUROC of 0.69 and an AUPRC of 0.58 when tested on the validation dataset for sub-challenge 1.

#### Team AI4knowledgeLAB

To predict the risk of PTB, a workflow based on an ensemble of random forest^43^ models with oversampling of the minority class had been used. For the implementation of the model, both metadata and characteristic data of the vaginal microbiome were used. Concerning metadata, information on race and ethnicity and the gestational week when the sample was collected were included into the analysis. Microbiome data included: relative abundances of clusters of variants measured at three different phylogenetic distances (0.1, 0.5, 1), alpha-diversity metrics, and “VALENCIA Community State Types” (CST). The pipeline is shown in Supplementary Figure 7.

The first step was to eliminate samples collected after the 32nd week of gestation. A model was then built that takes three different matrices as input, one for each phylogenetic distance, to create three independent models that can output three different predictions for the same individual, which are then combined using an ensemble strategy. Each input matrix had a number of features of 9743, 3651, and 1871: to each matrix of relative abundance of phylotypes were added features related to: alpha-diversity (7), CST (11), and demographics (8).

To make the dataset more balanced, a data augmentation algorithm, SMOTE (Synthetic Minority Oversampling Technique)^44^, was adopted. As a classification algorithm, random forest was chosen using the default parameters of the Scikit-learn python package^40^ due to its efficiency in handling datasets with a high number of features^45^. The final output was obtained as the average of the three probability values and the associated class was obtained from the probability value by imposing the classic threshold of 0.5. The prediction model achieved an AUROC of 0.64 and an AUPRC of 0.48 on the Dream Challenge validation dataset.

### Sub-challenge 2 - Top performing Teams

#### Team Techtmann Lab

To predict early PTB, a basic random forest classifier was employed using python’s Scikit-learn package^40^. Training data included relative abundances clustered phylogenetically at a distance of 0.1, race of the patient, VALENCIA community state types, diversity metrics, and collection week. This model used default Scikit-learn parameters and involved no additional feature selection or hyperparameter tuning. When tested on the competition validation dataset, the model reported an AUROC of 0.87 and an AUPRC of 0.45.

When investigating feature importance diversity metrics, race, community state type, sample collection week, and some phylotypes were found to be the most important features in the model’s decision-making. Specifically, five phylotypes whose relative abundances were identified as important to predict early PTB: *Lactobacillus jensenii, Lactobacillus iners, Lactobacillus crispatus, Prevotella bivia, and Ureaplasma urealyticum*. This approach is hypothesized to result in a model that was not over-tuned to the training data, allowing it to generalize well to the competition validation dataset.

#### Team KBJ

With the approach of team KBJ for sub-challenge 2, several processes were applied to improve the model prediction performance (Supplementary Figure 8). First, samples were filtered out by collection week conditions as the test dataset and aggregated all corresponding features. Here, one feature type was selected among several for taxonomy and phylotypes – genus-level and 0.1 phylogenetic distance, respectively. Also, race information was considered, while pairwise distance was excluded. Next, significant features were selected using the minimum redundancy maximum relevance^46^, which considers mutual information of features in terms of response variables (i.e., early preterm versus non-preterm). The feature selection was conducted for phylotypes, sequence variants, and taxonomy whose dimensions are relatively large compared to the data size. Then, an ensemble model was constructed with five algorithms (Linear Support Vector Classification^47^, Support Vector Classification^47^, Quadratic Discriminant Analysis^48^, Calibrated Classifier^49^, and Passive Aggressive Classifier^50^) that solely performed the best in crossvalidation. All compared models were tested with default parameters by the Lazy Predict^51^ and Scikit-learn^40^ python packages. The prediction model constructed by team KBJ achieved an AUROC of 0.841 and an AUPRC of 0.270 on the Dream Challenge validation dataset. Specifically, the model showed good balanced accuracy (sensitivity: 0.77; specificity: 0.79).

#### Sensitivity analysis on gestational age at sampling

To ensure that the best performing models were not overly reliant upon the gestational week of collection of specimens, we performed a sensitivity analysis–removing gestational age at sampling or permuting gestational age values (Table 5). Model performance was only modestly affected removing model access to the gestational age of collection, indicating the predictions were primarily based on other attributes.

**Table 5:**
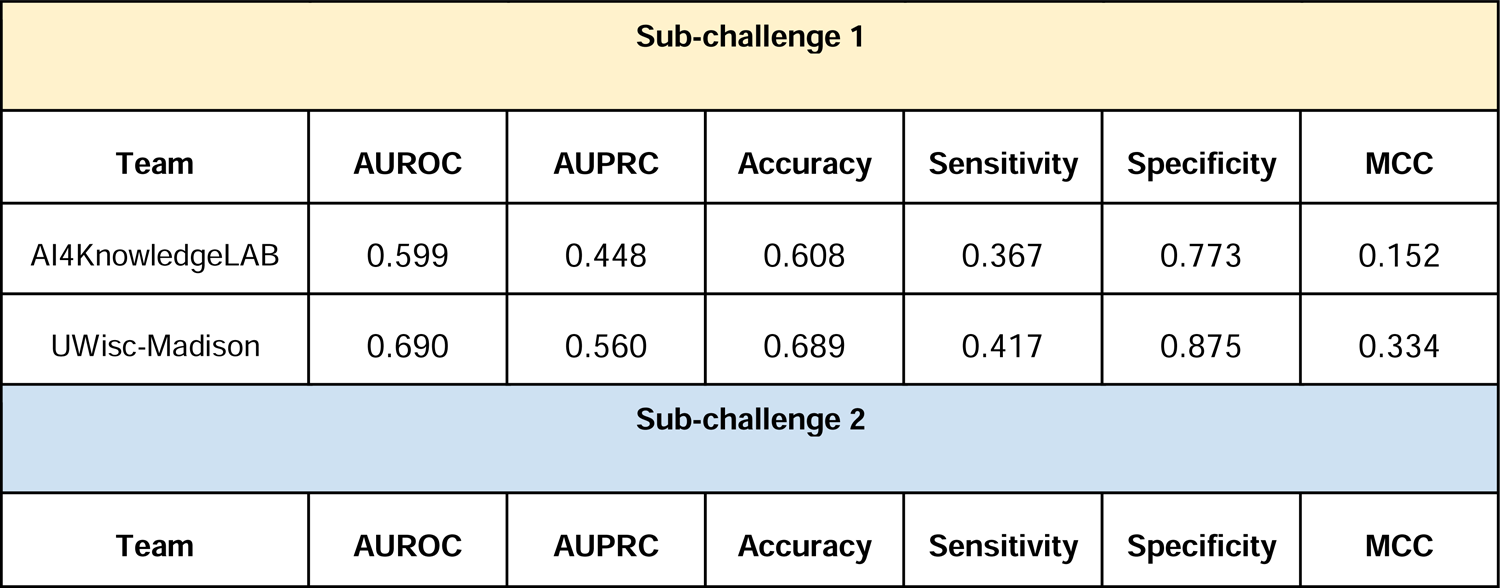
Sensitivity analysis removing gestational age as a feature for sub-challenge 1 and sub-challenge 2

#### Post-challenge ensemble models

Several ensemble models were created - combining results of (a) the winning teams, (b) the teams with Bayes factor < 20 (Tables 3 and 4), and (c) all the participants across the two sub-challenges (Figure 7). An improvement in performance was observed across the board with the ensemble models of Bayes factor < 20 performing the best AUROC 0.74 and AUROC 0.91 respectively for sub-challenges 1 and 2.

**Figure 7:**
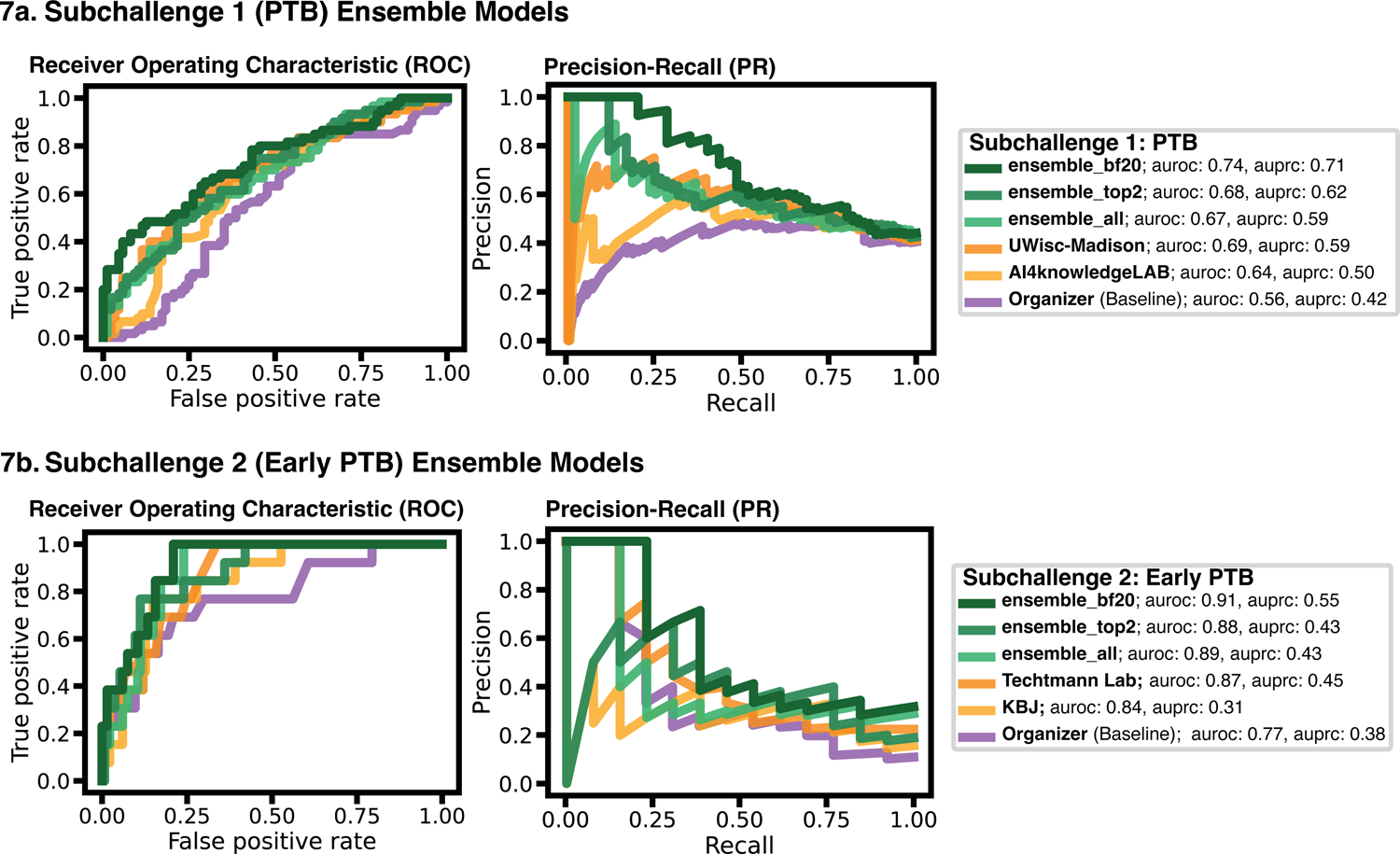
Ensemble Model Results. For a) sub-challenge 1 and b) sub-challenge 2, the area under the receiver operator characteristics (AUROC, left) curve and area under the precision-recall curve (AUPRC, right) of three ensemble models (‘ensemble_top2’: top two performing models, ‘ensemble_top2’: models with Bayes factor less than 20; and ‘ensemble_all’: all models), as well as first place, second place, and baseline models, colored by model

## Discussion

PTB, particularly early PTB (before 32 weeks of gestation), remains a potentially devastating outcome of pregnancy. Without a clear way of identifying pregnancies at risk for PTB, it remains difficult to target interventions or clinical trials. The microbiome has been extensively correlated in single-center studies with the risk for PTB, opening the promise of using the vaginal microbiome to build rigorous, generalizable, and robust predictive models to identify pregnancies at risk for PTB. However, results from various studies were largely inconclusive. In addition, combining data from different microbiome studies into a predictive, stable, and generalizable set of features for the rigorous evaluation of predictive models against independent validation datasets and their eventual use with vaginal microbiome data from individual pregnancies clinically is non-trivial. In this study, we leveraged data from 9 independent studies of the vaginal microbiome during pregnancy. The data was aggregated from public domain sources including dbGAP and the MOD Database for Preterm Birth Research. The final training dataset included data from 3,578 samples across 1,268 individuals, with 851 individuals delivering at term and 417 delivering preterm, including 170 early preterm deliveries. We applied a novel scientific and technical schema (implemented in a software workflow MaLiAmPi) for harmonizing microbiome data at the sequence-level, even when generated with different underlying primers and sequencing platforms, to transform the raw data into a stable and generalizable set of features suitable for predictive modeling. This schema also allowed the *post hoc* integration of microbiome data from two independent validation sets (that were unavailable at the time of the release of the training set) into the same set of features: an unpublished dataset from Wayne State University/Perinatology Research Branch and a second validation dataset generated by UCSF from samples provided by Stanford University. Crowdsourced predictive models were developed by 318 teams based on the training feature set and evaluated against the independent validation data within the same schema of features. Multiple teams were able to generate predictive models for both PTB and early PTB, with the models predicting the latter particularly robustly.

We noted that the best-performing predictive models all employed some type of feature-pruning and selection, typically within the broad family of random forest-like models. Given the sparseness of microbiome data, and plethora of features that can be detected, it is not surprising that modeling techniques more resilient to overfitting, and better able to hone in on the most important features, performed better. This risk of overfitting also speaks more broadly to the value of validating microbiome associations and predictive models on independent data sets; even with a large training data set consisting of multiple studies, teams often needed to adjust their models to reduce the risk of overfitting to perform well against the validation data.

While taxon data were provided to teams (the current state of the art for combining microbiome data), it is notable that the novel taxonomy-independent phylotypes were used by a majority of the better performing models. Taxonomy-based features were overall a challenge for participants, as there was poor overlap between the training and validation data sets at the taxonomic level. This required teams that relied upon taxonomy to preprocess the taxonomic feature tables, drop taxa that were not observed in the training data. In contrast, the taxonomy-independent phylotypes were intrinsically overlapped between the training and validation data.

An expected finding in our study is that more severe cases which involve early delivery were easier to predict from vaginal microbiome data than all PTB cases. This result was consistent for multiple independent modeling teams, including teams who tackled both sub-challenges, with sub-challenge 2 (predicting early PTB) models generating better predictions (as judged by our metrics, including AUROC). This was expected given that in early PTB the frequency of intra-amniotic infection is higher, and infection may be caused by ascending bacteria it’s been previously observed that there is a relationship between PTB and the vaginal microbiome^52^ also, it’s potentially a more extreme phenotype (rephrase).. Further study is needed, but we believe this could suggest that the vaginal microbiome has a stronger causal contribution to early PTB.

Through feature-permutation combined with multiple independently-developed highly-predictive models, we were able to identify multiple organisms, community state types, and community structures that associate with the risk for PTB opening the door to future studies into the underlying biology and pathophysiology of these associations, as well as more precise and effective intervention upon the vaginal microbiome during pregnancy to reduce the risk of PTB. In particular, while non-dominance of *Lactobacillus* in vaginal microbiome communities has previously been associated with PTB^26–28^, there seems to be physiologicallyrelevant species- and strain-level variability within the *Lactobacillus* and *Gardnerella* genera across pregnancy trimesters that deserves further exploration and indicates a potential role forintra-niche competition in the vaginal microbiome during pregnancy and the risk for early PTB.

The training data set itself, inclusive of the stable and generalizable features, is an invaluable resource for future studies of the vaginal microbiome during pregnancy. This training set, and more importantly the stable set of features, is a possible means of avoiding a challenge in the microbiome literature, where each study reports on a slightly different set of features. Future novel studies can leverage this large, geographically diverse, and strictly formatted data set to leverage and validate their findings.

The study has several limitations that should be considered when interpreting the results. The study is based on publicly available data which might not have full clinical or demographic annotations of the samples in the metadata. In particular, the spontaneous nature of PTB could not be ascertained for all patients in the training set, and differentiating between spontaneous preterm labor and delivery and preterm prelabor rupture of the membranes was not feasible. Recent work suggests that this latter phenotype is more likely associated with the microbiome^53, 54^. While the sample size of the study is considerable, with 3,578 samples across 1,268 individuals, it may not be representative of the entire population of pregnant women from around the world. We only considered binary outcomes (term vs preterm delivery) and did not take into account other important outcomes such as low birth weight or neonatal morbidity. The study is a computational challenge, and the results of the models are only as good as the data that they are trained on, and the limitations of the data may be reflected in the results. Finally, we only used data from the vaginal microbiome, which may not fully reflect the overall health of the pregnant women; other factors such as genetics, host-response, lifestyle, or environment may also play a significant role in parturition timing.

This work serves as the basis for several potential follow up studies. To improve the performance of the models, additional data such as demographic, clinical data, environmental data, or data from other body sites could be incorporated into the models. To better understand the mechanisms underlying PTB and early PTB further in vitro and in vivo validation of individual microbial features identified by the models can point to the underlying molecular mechanisms of human parturition. Studying how to in turn modulate the microbes can result in therapeutic hypotheses. Once the models have been validated and optimized, the next logical step is to translate them into clinical practice to help identify women at risk of PTB and to develop appropriate interventions to prevent PTB.

Together, we believe this represents a genuine advancement in our ability to identify pregnancies at risk for early PTB. Given these models rely upon a generalizable set of features that can accommodate post-hoc data from individual pregnancies, these predictive models are ‘shovel ready’ for use in clinical trials and exploration of their potential role in the clinical care of pregnancies. Further, we believe this scientific and technical schema could be suitable for building microbiome-based predictive models for other microbiomerelated conditions.

## Methods

Collection, generation, and analysis of vaginal microbiome data was approved by the National Heart, Lung, and Blood Institute (NHLBI) Clinical Data Science Institutional Review Board (CDS-IRB) in study number 2021-040, and reliance was granted to the NHLBI CDS-IRB by the University of California, San Francisco Institutional Review Board in study number 21-35274.

### Training Data Acquisition and Processing

The following vaginal microbiome studies were identified by leveraging the March of Dimes Preterm Birth database^38^, the NCBI Sequence Read Archive^55^, the European Nucleotide Archive^56^, and the database of Genotypes and Phenotypes (dbGaP)^37^. Sequence data and associated metadata for the DiGiulio et al.^27^ cohort were downloaded from ImmPort^57^, under Study SDY465 in May 2016. Sequence data and associated metadata for Romero et al.^58^ cohort were downloaded from the NCBI Sequence Read Archive under BioProject PRJNA242473 in May 2016. Sequence data and associated metadata for the Callahan et al.^28^ cohort were downloaded from the NCBI Sequence Read Archive under BioProject PRJNA393472 in January 2018. Sequence data and associated metadata for the Stout et al.^59^ cohort were downloaded from the NCBI Sequence Read Archive under BioProject PRJNA294119 in January 2018. Sequence data for the Kindinger et al.^60^ cohort were downloaded from the Sequence Read Archive of the European Nucleotide Archive under Projects PRJEB11895 and PRJEB12577 in June 2020, and associated metadata was downloaded from Additional Files 4 and 6 from the paper with some additional metadata requested from the senior author. Sequence data and associated metadata for the Brown et al. (2018)^61^ cohort were downloaded from the Sequence Read Archive of the European Nucleotide Archive under Project PRJEB21325 in June 2020 with some additional metadata requested from the senior author. Sequence data and associated metadata for the Brown et al. (2019)^53^ cohort were downloaded from the Sequence Read Archive of the European Nucleotide Archive under Project PRJEB30642 in June 2020 with some additional metadata requested from the senior author. Sequence data and associated metadata for the Elovitz et al.^62^ cohort were downloaded from the database of Genotypes and Phenotypes (dbGaP)^37^ under accession number phs001739.v1.p1 in September 2021. Sequence data and associated metadata for the Fettweis et al.^63^ cohort were downloaded from the NCBI Sequence Read Archive under BioProject ID PRJNA430482 in January 2022, and associated metadata were requested through and obtained from the RAMS Registry (https://ramsregistry.vcu.edu).

### Validation Data Generation

#### Wayne State University

##### Study design, sample collection

The microbiome dataset from Wayne State University School of Medicine included in the challenge was a subset of randomly selected 20 cases and 40 controls from a larger retrospective longitudinal case-control study described in detail elsewhere (https://www.researchsquare.com/article/rs-2359402/v1)^54^. The 20 spontaneous PTB cases included both spontaneous preterm labor with intact membranes (PTL) and preterm prelabor rupture of membranes (PPROM) resulting in delivery 20-36+6 weeks. Cases had 3 or 4 longitudinal samples collected from 10-36 weeks of gestation which were matched with samples from controls (2 to 4 samples per patient). Term controls were defined as women who delivered between 38 and 42 weeks of gestation without congenital anomalies or obstetrical, medical, or surgical complications. Samples of vaginal fluid were collected using a Dacron swab (Medical Packaging Corp., Camarillo, CA). Vaginal swabs were stored at −80°C until time of DNA extraction, following established standard operating procedures. The study was conducted at the Perinatology Research Branch, an intramural program of the Eunice Kennedy Shriver National Institute of Child Health and Human Development, National Institutes of Health, U.S. Department of Health and Human Services, Wayne State University (Detroit, MI), and the Detroit Medical Center (Detroit, MI). The collection of samples was approved by the Institutional Review Boards of the National Institute of Child Health and Human Development and Wayne State University (#110605MP2F(RCR)). All participating women provided written informed consent prior to sample collection.

##### DNA extraction from vaginal swabs

Genomic DNA was extracted from vaginal swabs using a Qiagen MagAttract PowerMicrobiome DNA/RNA EP extraction kit (Qiagen, Germantown, MD), with minor modifications to the manufacturer’s protocols as described in (https://www.researchsquare.com/article/rs-2359402/v1). The purified DNA was transferred to the provided 96-well microplates and stored at −20°C.

##### 16S rRNA gene sequencing and processing

The V4 region of the 16S rRNA gene was amplified from vaginal swab and control DNA extracts and sequenced at Michigan State University’s Research Technology Support Facility (https://rtsf.natsci.msu.edu/) using the dual indexing sequencing strategy developed by Kozich et al.^64^. The forward primer was 515F: 5’-GTGCCAGCMGCCGCGGTAA-3’ and the reverse primer was 806R: 5’GGACTACHVGGGTWTCTAAT-3’.

### Stanford University

#### Study design, sample collection

The Stanford University microbiome dataset included in the challenge consisted of 40 cases and 48 controls from a repository of specimens from women enrolled in a longitudinal study conducted by the March of Dimes Prematurity Research Center at Stanford University. Samples of vaginal fluid were collected using a 2x Sterile Catch-All™ Sample Collection Swab (Epicentre Biotechnologies #QEC091H, Madison, WI). Vaginal swabs were placed into tubes then immediately placed on ice or in a household freezer (−20°C). After samples arrived at the March of Dimes Prematurity Center they were immediately placed on dry ice, inventoried, and then stored at −80°C at the Stevenson Laboratory until time of DNA extraction. The study was conducted at Stanford Hospital and Clinics. The collection of samples was approved by the Institutional Review Board of Stanford University (Study number 21956). All participating women provided written informed consent prior to sample collection.

#### Vaginal swab DNA extraction and 16S rRNA sequencing

Genomic DNA extraction and microbial sequencing were performed at the Microbial Genomics CoLab Plugin Facility within the Benioff Center for Microbiome Medicine at University of California, San Francisco. First, vaginal swabs were aseptically transferred to 2 mL tubes pre-filled with 300 µL sterile molecular-grade water. Vaginal samples were vortexed with the swab remaining in the tube. 200 μL vaginal suspension from the tube was withdrawn for downstream processing using the QIAamp BiOstic DNA Kit (QIAGEN, Hilden, Germany). DNA from all samples and several extraction blanks were extracted according to the manufacturer’s protocol and eluted in 50 µl EB buffer. DNA concentrations were quantified using the Qubit dsDNA HS Assay Kit (ThermoFisher Scientific, MA), diluted to 5 ng/µL and stored at −20°C.

The V4 hypervariable region of the 16S rRNA gene was amplified using 515F and 806R primers65 with PCR conditions previously described66. Amplicon reactions were quantified using the Qubit dsDNA HS Assay Kit (ThermoFisher Scientific, MA), and pooled at equimolar concentrations. The pooled library was cleaned and concentrated using the Agencourt AMPure XP beads (Beckman-Coulter), quality checked with the Bioanalyzer DNA 1000 Kit (Agilent, Santa Clara, CA), quantified using the KAPA Library Quantification Kit (KAPA Biosystems), and diluted to 2 nM. Library was denatured according to manufacturer’s protocol and spiked in with 40% PhiX control prior to loading onto the NextSeq 550 platform (Illumina, San Diego, CA) for 2 x 150bp sequencing.

#### Data Processing and Normalization

We applied MaLiAmPi^39^ to both training and test data to process and aggregate the datasets. Standardized processed data format facilitates running Docker containers, as we had participants use in our Challenge, and choosing feature sets for permutation. MaLiAmPi is available as a nextflow workflow, and is 100% containerized to allow for usage on multiple different high performance computing resources. In brief, MaLiAmPi uses DADA2 to assemble each project’s raw reads into approximate sequence variants (ASVs). These ASVs are used to recruit full-length 16s rRNA gene alleles from a repository (based on sequence identity). These recruits are assembled into a de novo maximum-likelihood phylogeny with RAxML and the ASVs are placed onto this common phylogenetic tree with EPA-ng. Finally, these placements are used to determine the alpha-diversity of communities (diversity measures include Shannon, Inverse Simpson, Balance weighted phylogenetic diversity (bwpd), phylogenetic entropy, quadratic, unrooted phylogenetic diversity, and rooted phylogenetic diversity), phylogenetic (KR) distance between communities, provide taxonomic assignments to each ASV, and cluster ASVs into phylotypes (based on phylogenetic distance between ASVs). Sequence variance counts were also determined. In addition, VALENCIA^67^ was used to provide the community state type (CST) of each sample and alluvial plots were made using the ggalluvial R package^68^ in order to visualize CST composition by trimester. UMAP representations of the data and violin plots of Shannon alpha diversity before and after processing of the data with MaLiAmPi were visualized to gauge data harmonization. Extensive use of the Python seaborn visualization package was used for figure preparation.

### DREAM Challenge

#### Overall Challenge structure

The overview of the Challenge is shown in Figure 1. All Challenge elements were supported by the Synapse platform (http://www.synapse.org), including documentation, access to the data, submission of models, leaderboards, and the discussion forum. To gain access to the data, teams were required to comply with a data use agreement, restricting use of the data outside the Challenge and providing guidelines on ethical participation in the Challenge. Teams were provided the training data, they built their models, dockerized their environment, and submitted their models through the Synapse platform. Models were run on the test data and performance metrics were returned to the teams. Teams were limited to 5 total submissions with the top performing model selected as the final submission to be scored and ranked. Leaderboards were provided throughout the open phase of the Challenge, which provided teams with real-time feedback and comparative performance rankings. After the close of the Challenge, models were evaluated for completeness and reproducibility. For teams to be included in the Preterm Birth DREAM Community, they were required to make the code public, provide a method write-up, and participate in a post-challenge survey to collect information on method development and features of the data important to the model.

#### Participant engagement

Information about our challenge was shared through the Dream Challenges website (https://dreamchallenges.org). Challenge organizers also shared information about the challenge through listservs such as ML-news Google News Group and social media outlets including Facebook, LinkedIn, Reddit, and Twitter.

In order to preserve model environments for portability of models, we required participants to submit Docker environments. These environments contain the necessary programming dependencies and models for each sub-challenge that can run on a processed and prepared microbiome dataset folder arranged in a standardized format. The organizers prepared an example Docker container for participants to utilize as a starting template and held occasional seminars to describe the data and answer questions from participants. Organizers also engaged with participants through the forums to help answer questions throughout the challenge.

#### Assessment Strategies

Performance metrics that were used to evaluate the teams include Area under the receiver operator characteristic (AUROC) curve and Area under the precision-recall (AUPR) curve. On the held-out external validation dataset, metrics of accuracy, sensitivity, and specificity were also computed. These metrics were shown on the final public rankings.

The reproducibility of models, including the baseline, were determined by calculating the Bayes factor for 1000 bootstrapped iterations on a random sampling of the data. For each sub-challenge, the best-performing models from each team were rerun to obtain scores on the random sampling. These scores were then used to calculate the Bayes factor, using the computeBayesFactor function from the challenge scoring R package^69^, comparing them to the top-performing model as well as the baseline model.

To increase our certainty of DREAM Challenge participants’ rankings whose models’ performances could have been affected by prediction threshold and class imbalance in our validation dataset, we employed the following strategies to validate participants’ models for both sub-challenges on the external dataset: inverting labels, bootstrapped random subsampling, bootstrapped under-sampling, and bootstrapped over-sampling.

### Inverted labels

Invert the class labels for the external dataset and prediction model outputs (i.e., classifying preterm or early preterm births as term births, and vice versa), and computing AUROC/AUPR curves.

Bootstrapped random subsampling:

Randomly sample a subset of 100 from the 152 participants of the external dataset, and run the prediction models on the validation data subset, bootstrapped 1000 times.

Bootstrapped undersampling:

Undersample the external dataset (n = 152) to balance the minority (Preterm, n = 63. Early preterm, n=13) and majority (i.e., Term, n = 89) classes by randomly sampling from the minority and the majority groups to have the same number in each group (n = 50 for Preterm and n = 50 for Term in sub-challenge 1, and n = 13 for Early Preterm and n = 13 for Term in for sub-challenge 2), and then computing AUROC/AUPRC on the undersampled external validation dataset, bootstrapped 1000 times.

Bootstrapped oversampling:

Oversample the external dataset to balance the preterm or early preterm and term classes by randomly sampling per group (n = 200 for Preterm and n = 200 for Term in sub-challenge 1, and n = 200 for Early Preterm and n = 200 for Term in for sub-challenge 2), and then computing AUROC/AUPRC oversampled external dataset, bootstrapped 1000 times.

Individual team methods are linked to in Supplementary Table 1.

DREAM challenge participants and teams were surveyed to gather information on how they developed their models.

Feature importance was determined across the best performing models for sub-challenges 1 and 2 that demonstrated predictive performance at threshold of 0.64 for sub-challenge 1 and a threshold of 0.80 subchallenge 2 which also could be run in a bootstrapped manner in a tractable amount of time

Sensitivity analysis was carried out removing gestational age at sampling as a feature.

As with previous DREAM Challenges, ensemble models were generated to explore the “wisdom of the crowds” phenomenon, by aggregating the best-performing models from each team. For each sub-challenge, we experimented with 3 ensemble models by calculating the mean estimation from: 1) top two performing models; 2) models with Bayes factor less than 20; 3) all models.

## Supporting information

Combined Supplements

## Author contributions

JG, TTO, AT, AR, IK, GA, ALT, JC, and MS conceived the study. CWYH, RJW, and ALT generated and shared data for the validation dataset. JG, TTO, AST, AR, and SSM aggregated the training datasets. JG and AR normalized the training and validation datasets. JG, TTO, AST, AR, VC, ALT, JC, and MS analyzed and interpreted the data regarding the vaginal microbiome, the DREAM challenge outcomes, and findings from the challenge participants models. JG, TTO, AST, CWYH, RJW, ZW, PN, AK, EK, ALT, JC, and MS were major contributors in writing the manuscript. All authors read and approved the final manuscript.

## Data availability

Sequence data and associated metadata for Study SDY465 were downloaded from ImmPort^57^ via the March of Dimes Preterm Birth database^38^. Sequence data and associated metadata for BioProjects PRJNA242473, PRJNA294119, PRJNA393472, and PRJNA430482 were downloaded from the NCBI Sequence Read Archive^55^. Additional associated metadata for PRJNA430482 were requested through and obtained from the RAMS Registry (https://ramsregistry.vcu.edu).

Sequence data and associated metadata for Projects PRJEB11895, PRJEB12577, PRJEB21325, and PRJEB30642 were downloaded from the Sequence Read Archive of the European Nucleotide Archive^56^, with associated metadata for PRJEB11895 and PRJEB12577 downloaded from Additional Files 4 and 6 from the paper by the Kindinger et al.^60^. Additional associated metadata for Projects PRJEB11895, PRJEB12577, PRJEB21325, and PRJEB30642 were requested from the senior author.

Sequence data and associated metadata for accession number phs001739.v1.p1 were downloaded from the database of Genotypes and Phenotypes (dbGaP)^37^.

The training dataset representing 7 of the 9 aggregated studies and the validation dataset for our Challenge are available under Study ID SDY2187 from the MOD Preterm Birth Research Database (https://pretermbirthdb.org/mod/studydata). Two of the nine training data (PRJNA430482 and phs001739.v1.p1.) are exclusively available via dbGap after following the application procedures there.

## Code availability

The code for the microbiome data harmonization tool, MaLiAmPi, is available at https://github.com/jgolob/maliampi. DREAM challenge participants’ code for sub-challenge 1 and sub-challenge 2 is in their docker submissions which may be accessed by the hyperlinks listed in Supplemental Tables 1 and 2, respectively, of this work.

## Acknowledgements

We thank members of the Sirota Lab, University of California, San Francisco, for useful discussion. This study was supported by the March of Dimes (JLG, TTO, AR, AST, VC, CWYH, RJW, KF, GA, IK, JB, AN, JG, ZW, PN, AK, IB, EK, SJ, SN, YSLL, PRB, DAM, SVL, JA, DKS, NA, JCC, MS) and R35GM138353 (NA), 1R01HL139844 (NA), 3P30AG066515 (NA), 1R61NS114926 (NA), 1R01AG058417 (NA), R01HD105256 (NA, MS), P01HD106414 (NA), the Burroughs Welcome Fund (NA), the Alfred E. Mann Foundation (NA), and the Robertson foundation (NA), Spanish Ministry of Science, Innovation and Universities through FPU program FPU18/0177; EST22/00170 (ALP), Instituto de Salud Carlos III (Spanish Ministry of Science and Innovation) through Miguel Servet program CP20/00118 and co-funded by European Union (PGD).

## FIGURE LEGENDS

Supplementary Figure 1: <u>Individual study designs</u>. Gestational week at sample collection stratified by study and colored by birth outcome

Supplementary Figure 2: <u>UMAP ordination plots</u> of data after harmonization, where each dot represents one vaginal microbiome sample, colored a) by trimester and b) by race/ethnicity

Supplementary Figure 3: <u>CST heatmap</u>. Heatmap of community state types (CST) for term, preterm, and early preterm births across the first, second, and third trimesters of pregnancy

Supplementary Figure 4: <u>Bootstrapped results for sub-challenge 1: preterm birth prediction.</u> Top includes curves for inverted labels in the test set, while bottom includes undersampling and oversampling per group (preterm/term) to ensure balance between groups. Left is AUROC, right is AUPRC

Supplementary Figure 5: <u>Bootstrapped results sub-challenge 2: early preterm birth prediction.</u> Top includes curves for inverted labels in test set, while bottom includes undersampling and oversampling per group (early preterm/not early preterm) to ensure balance between groups. Left is AUROC, right is

### AUPRC

Supplementary Figure 6: <u>Overview of the pipeline of U-Wisconsin team.</u> The architecture of the pipeline for prediction of preterm birth using microbiome data and metadata. CLR is applied to each type of the microbiome count data. Rare microbial features are filtered out. Two LightGBM models are trained: one on all available specimen data (Model 1), and another on data from Project G only (Model 2). The predictions from these models are then combined, and the aggregate prediction is used to generate a probabilistic prediction of preterm birth

Supplementary Figure 7: <u>Workflow of the analysis by Team AI4knowledgeLAB.</u> The probability score of the final output was obtained as the average of the 3 probability values and the associated class was obtained from the probability value by imposing the classic threshold of 0.5

Supplementary Figure 8: <u>Overview of the model of Team KBJ.</u> Left represents preprocessing of provided metadata and processed outputs from MaliAmPi pipeline.They extracted samples according to the test set condition and aggregated features to represent participants. Then, sparse feature types were handled with mRMR and concatenated with other features. Additional race information was used as a feature. For ensemble learning, based on 26 different machine learning models, five algorithms were selected by topranked models. The final predicted value was calculated as the mean of each probability

Supplementary Figure 9: Features of sub-challenge 1 for top model for top teams with threshold or above baseline (same criteria as figure 5)

Supplementary Figure 10: Features of sub-challenge 2 for top model for top teams with threshold or above baseline (same criteria as figure 5)

## The Preterm Birth DREAM Community

Yong Ju Ahn^1^, Yadid M. Algavi^2^, Nicola Amoroso^3,4^, Maria De Angelis^5^, George Austin^6,7^, Ashley Babjac^8^, Daehun Bae^9^, Seungheun Baek^10^, Roberto Bellotti^4,11^, Panayiotis Benos^12^, Yonatan Berg^2^, Isaac Bigcraft^13^, Aya Brown-Kav^6^, Kun Bu^14^, Guanhua Chen^15^, Jhih-Yu Chen^16^, Sz-Hau Chen^17^, Tsai-Min Chen^18,19^, Feng Cheng^20,21^, Junseok Choe^10^, Francesco Cremonesi^22^, Saishi Cui^23^, Yang Dai^24^, Scott Emrich^8^, Alonso Felipe-Ruiz^25^, Diego Fernandez-Edreira^26,27^, Carlos Fernandez-Lozano^26,27^, Jifan Gao^15^, Sergio Pérez García^28^, Mogan Gim^10^, Enrico Glaab^29^, Akhil Goel^30^, Ella Goldschmidt^31^, Igor Goryanin^32,33^, Yuanfang Guan^34^, Dror Hadas^31^, Kyudong Han^35^, Weiru Han^14^, Chih-Han Huang^36^, Kuei-Lin Huang^37^, Hirotaka Iijima^38,39^, Gwanghoon Jang^10^, Jongbum Jeon^40^, Hongmei Jiang^41^, Michael Jochum^42^, Soobok Joe^40^, Jaewoo Kang^10,43^, Tina Khajeh^24^, Eunyoung Kim^9^, Hajung Kim^43^, Jiwoong Kim^44,45^, William Kindschuh^6^, Stephanie Kivlin^46^, Hayata Kodama^38^, Aki Koivu^47^, Tal Korem^6,48^, Abigail Kuntzleman^13^, Manuel E. González Lastre^49^, Mo Li^50^, Jose Linares-Blanco^51,52^, Wodan Ling^53^, Tyler C Lovelace^54,55^, Jiuyao Lu^50^, Zhixiu Lu^8^, Jiangyue Mao^30^, Miguel Pineda Martín^56^, Yusuke Matsui^38,57^, Kevin McPherson^58^, Alfonso Monaco^4,11^, Hesam Montazeri^59^, Chengcheng Mou^60^, Efrat Muller^31^, Akiha Nakano^61^, Saina Nassiri^62^, Sina Nassiri^63^, Naresh Nelaturi^64^, Milad Norouzi^59^, Pierfrancesco Novielli^4,5^, Indumathi Palanikumar^65,66,67^, Tanay Panja^68^, Ester Pantaleo^4,11^, Itsik Pe’er^6,7^, Omri Peleg^31^, Anna Plantinga^69^, Augustinas Prusokas^58^, Alisa Prusokiene^70^, Karthik Raman^65,66,67^, Derek Reiman^71^, Renata Retkute^72^, Donato Romano^4,5^, Gail Rosen^73^, Mohammad Sadegh Vafaei Sadi^59^, Mikko Sairanen^47^, Hibiki Sakata^38^, Ricardo Paixao dos Santos^74^, Edward S.C. Shih^75^, Koji Shimazaki^38^, Guy Shur^2^, Alireza Fotuhi Siahpirani^59^, Vijaya Yuvaram Singh V M^66,67,76^, Himanshu Sinha^65,66,67^, Bahrad Sokhansanj^73^, Anatoly Sorokin^32^, Go Suhara^61^, Zheng-Zheng Tang^15^, Sabina Tangaro^4,5^, Victor Tarca^77^, Stephen Techtmann^13^, Manoj Teltumbade^64^, Ambuj Tewari^30^, Gabriel Trigo^7^, Mor Tsamir^2^, Kako Tsukioka^61^, Kohei Uno^38^, Mirco Vacca^5^, Hsuan-Kai Wang^58^, Huiqian Wang^30^, Zehua Wang^30^, Zijing Wang^78^, Zidan Wang^41^, Zhoujingpeng Wei^15^, Chih-Hsun Wu^79^, Michael C. Wu^80^, Shaoming Xiao^81^, Ryota Yanase^61^, Jiaming Yao^30^, Issa Zakeri^23^, Wenjie Zeng^12^, Xiaowei Zhan^44,45^, Liangliang Zhang^82^, Yuci Zhang^30^, Ni Zhao^50^

^1^Division of Microbiome, HuNBiome Co., Ltd., Seoul, Republic of Korea.

^2^Sackler Faculty of Medicine, Tel Aviv University, Tel Aviv, Israel.

^3^Dipartimento di Farmacia - Scienze del Farmaco, Universita degli Studi di Bari A. Moro, Bari, Italy.

^4^Istituto Nazionale di Fisica Nucleare, Sezione di Bari, Bari, Italy.

^5^Dipartimento di Scienze del Suolo, della Pianta e degli Alimenti, Universita degli Studi di Bari A. Moro, Bari, Italy.

^6^Program for Mathematical Genomics, Department of Systems Biology, Columbia University Irving Medical Center, New York, New York, United States.

^7^Department of Computer Science, Columbia University, New York, New York, United States.

^8^Department of Computer Science, Univeristy of Tennessee, Knoxville, Tennessee, United States.

^9^School of Electrical Engineering and Computer Science, Gwangju Institute of Science and Technology, Gwangju, Republic of Korea.

^10^Department of Computer Science and Engineering, Korea University, Seoul, Republic of Korea.

^11^Dipartimento Interateneo di Fisica M. Merlin, Universita degli Studi di Bari A. Moro, Bari, Italy.

^12^Epidemiology, University of Florida College of Public Health and Health Professions and College of Medicine, Gainesville, Florida, United States.

^13^Department of Biological Sciences, Michigan Technological University, Houghton, Michigan, United States.

^14^Department of Mathematics & Statistics, College of Art and Science, University of South Florida, Tampa, Florida, United States.

^15^Department of Biostatistics and Medical Informatics, University of Wisconsin-Madison, Madison, Wisconsin, United States.

^16^Graduate Institute of Biomedical Electronics and Bioinformatics, National Taiwan University, Taipei, Taiwan.

^17^Industrial Information Department, Development Center for Biotechnology, Taipei, Taiwan.

^18^Graduate Program of Data Science, National Taiwan University and Academia Sinica, Taipei, Taiwan.

^19^Research Center for Information Technology Innovation, Academia Sinica, Taipei, Taiwan.

^20^Department of Pharmaceutical Science, Taneja College of Pharmacy, University of South Florida, Tampa, Florida, United States.

^21^Department of Biostatistics & Epidemiology, College of Public Health, University of South Florida, Tampa, Florida, United States.

^22^Centre Inria d’Université Côte d’Azur, Centre Inria, Biot, Sophia Antipolis, France.

^23^Department of Epidemiology and Biostatistics, Drexel University, Philadelphia, Pennsylvania, United States.

^24^Department of Biomedical Engineering, University of Illinois Chicago, Chicago, Illinois, United States.

^25^Department of Genomics and Proteomics, Institute of Biomedicine of Valencia, Valencia, Spain.

^26^Department of Computer Science and Information Technologies, Universidade da Coruña, A Coruña, Spain.

^27^CITIC-Research Center of Information and Communication Technologies, Universidade da Coruña, A Coruña, Spain.

^28^Data Engineer, Damavis Studio, Palma de Mallorca, Spain.

^29^Luxembourg Centre for Systems Biomedicine, University of Luxembourg, Esch-sur-Alzette, Luxembourg.

^30^Department of Statistics, University of Michigan, Ann Arbor, Michigan, United States.

^31^Blavatnik School of Computer Science, Tel Aviv University, Tel Aviv, Israel.

^32^Biological Systems Unit, Okinawa Institute of Science and Technology, Onna, Okinawa, Japan.

^33^School of Informatics, University of Edinburgh, Edinburgh, Midlothian, United Kingdom.

^34^Department of Computational Medicine and Bioinformatics, University of Michigan, Ann Arbor, Michigan, United States.

^35^Department of Microbiology, Dankook University, Cheonan, Republic of Korea.

^36^Department of Product, ANIWARE, Taipei, Taiwan.

^37^School of Medicine, China Medical University, Taichung, Taiwan.

^38^Graduate School of Medicine, Nagoya University, Nagoya, Aichi, Japan.

^39^Institute for Advanced Research, Nagoya University, Nagoya, Aichi, Japan.

^40^Korea BioInformation Center, Korea Research Institute of Bioscience & Biotechnology, Daejeon, Republic of Korea.

^41^Department of Statistics and Data Science, Northwestern University, Evanston, Illinois, United States.

^42^Obstetrics and Gynecology, Baylor College of Medicine and Texas Children’s Hospital, Houston, Texas, United States.

^43^Interdisciplinary Graduate Program in Bioinformatics, Korea University, Seoul, Republic of Korea.

^44^Peter O’Donnell Jr. School of Public Health, University of Texas Southwestern Medical Center, Dallas, Texas, United States.

^45^Quantitative Biomedical Research Center, University of Texas Southwestern Medical Center, Dallas, Texas, United States.

^46^Department of Ecology, University of Tennessee, Knoxville, Tennessee, United States.

^47^Advanced Research and Technology, R&D PerkinElmer, Wallac Oy, Turku, Finland.

^48^Department of Obstetrics and Gynecology, Columbia University Irving Medical Center, New York, New York, United States.

^49^Department of Theoretical Condensed Matter Physics, Universidad Autónoma de Madrid, Madrid, Spain.

^50^Department of Biostatistics, Johns Hopkins University, Baltimore, Maryland, United States.

^51^Department of Statistics, University of Granada, Granada, Spain.

^52^GENYO. Centre for Genomics and Oncological Research: Pfizer, University of Granada, Granada, Spain.

^53^Biostatistics Division in the Population Health Sciences Department, Weill Cornell Medicine, New York, New York, United States.

^54^Department of Computational & Systems Biology, University of Pittsburgh, Pittsburgh, Pennsylvania, United States.

^55^Joint CMU-Pitt PhD Program in Computational Biology, Carnegie Mellon University and University of Pittsburgh, Pittsburgh, Pennsylvania, United States.

^56^Matematician, University of Seville, Seville, Spain.

^57^Institute for Glyco-core Research (iGCORE), Nagoya University, Nagoya, Aichi, Japan.

^58^Independent Researcher.

^59^Department of Bioinformatics, Institute of Biochemistry and Biophysics, University of Tehran, Tehran, Iran.

^60^Department of Computer Science and Engineering, College of Engineering, University of South Florida, Tampa, Florida, United States.

^61^School of Health Sciences, Nagoya University, Nagoya, Aichi, Japan.

^62^Department of Gynecology and Obstetrics, Tehran University of Medical Sciences, Tehran, Iran.

^63^Pharma Research and Early Development, Roche, Basel, Switzerland.

^64^CognitiveCare Inc, CognitiveCare Inc, Milpitas, California, United States.

^65^Department of Biotechnology, Bhupat and Jyoti Mehta School of Biosciences, Indian Institute of Technology Madras, Chennai, India.

^66^Centre for Integrative Biology and Systems Medicine, Indian Institute of Technology Madras, Chennai, India.

^67^Robert Bosch Centre for Data Science and Artificial Intelligence, Indian Institute of Technology Madras, Chennai, India.

^68^Huron High School, Huron High School, Ann Arbor, Michigan, United States.

^69^Department of Mathematics and Statistics, Williams College, Williamstown, Massachusetts, United States.

^70^School of Natural and Environmental Sciences, Newcastle University, Newcastle, United Kingdom.

^71^Toyota Technological Institute at Chicago, Toyota Technological Institute at Chicago, Chicago, Illinois, United States.

^72^Department of Plant Sciences, University of Cambridge, Cambridge, United Kingdom.

^73^Department of Electrical & Computer Engineering, Drexel University, Philadelphia, Pennsylvania, United States.

^74^Department of Medicine, Universidade de São Paulo, São Paulo, Brazil.

^75^Institute of Biomedical Sciences, Academia Sinica, Taipei, Taiwan.

^76^Department of Biotechnology, Indian Institute of Technology Madras, Chennai, India.

^77^Clague Middle School, Clague Middle School, Ann Arbor, Michigan, United States.

^78^Data Science Institute, Columbia University, New York, New York, United States.

^79^Artificial Intelligence and E-learning Center, National Chengchi University, Taipei, Taiwan.

^80^Public Health Sciences Division, Fred Hutchinson Cancer Research Center, Washington, Seattle, United States.

^81^School of Medicine, Johns Hopkins University, Baltimore, Maryland, United States.

^82^Department of Population and Quantitative Health Sciences, Case Western Reserve University, Cleveland, Ohio, United States.

