## Supplementary material for "Microbiome Preterm Birth DREAM Challenge: Crowdsourcing Machine Learning Approaches to Advance Preterm Birth Research": Combined Supplements

**Figure S1:** Specimens collected per pregnancy

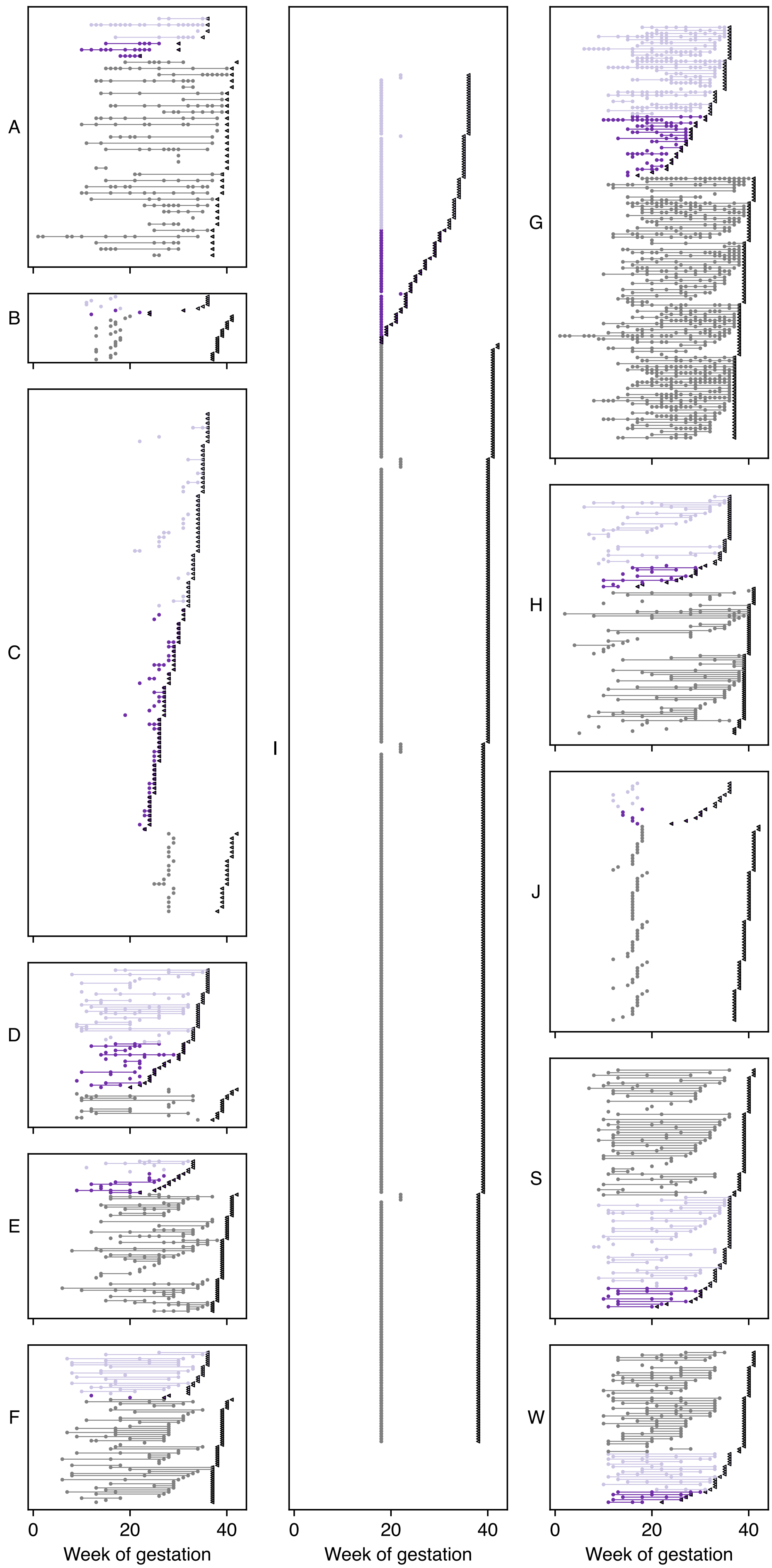

**Fig S2a: UMAP By Trimester** (Bray Curtis)

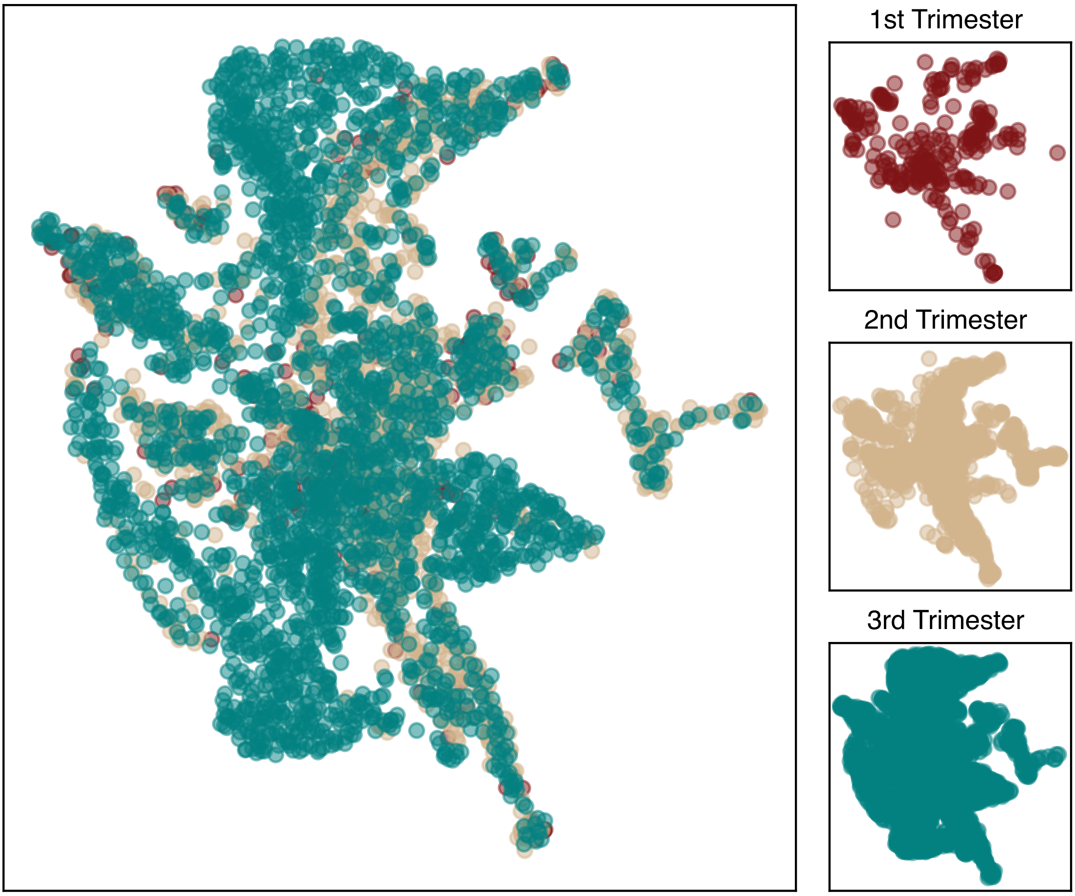


**Fig S2b: UMAP By NIH Racial Category** (Bray Curtis)

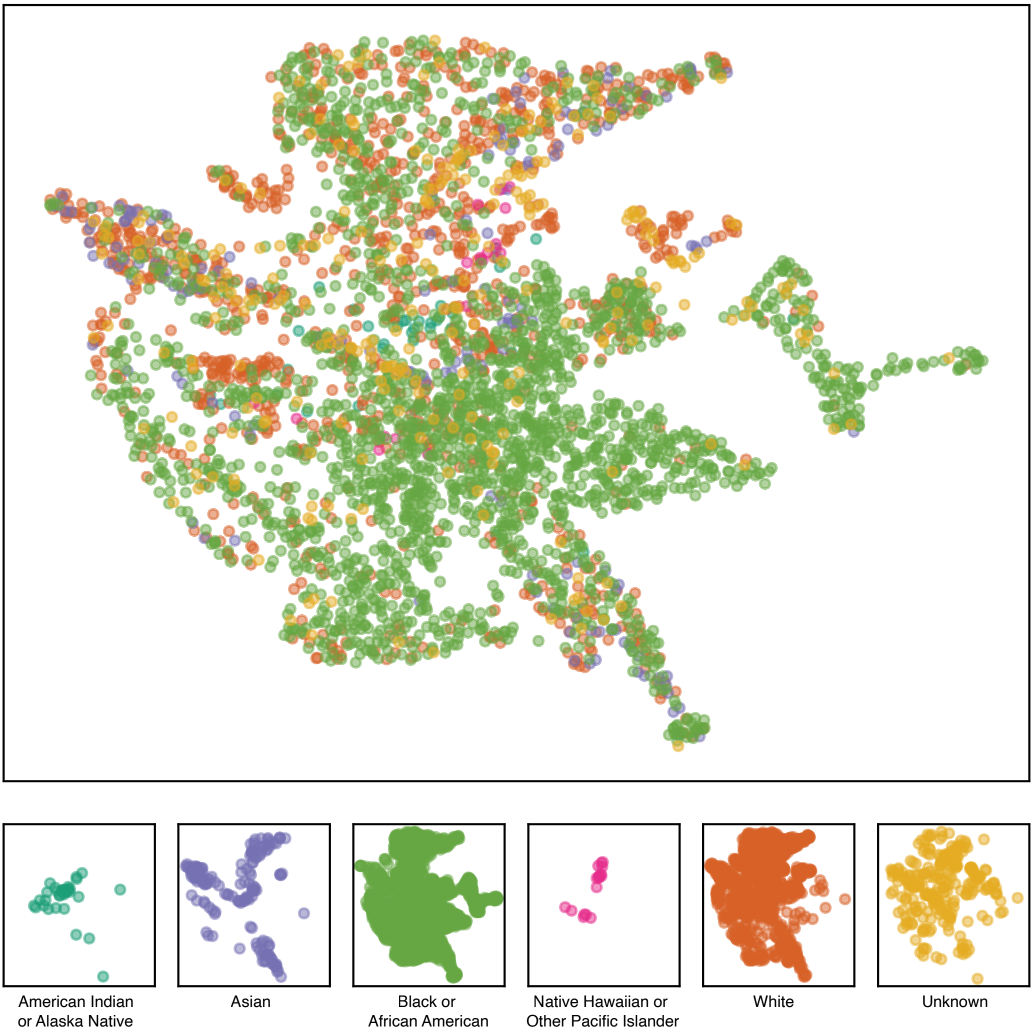

**Figure S3: Valencia Community State Type (CST) by Trimester and Outcome**

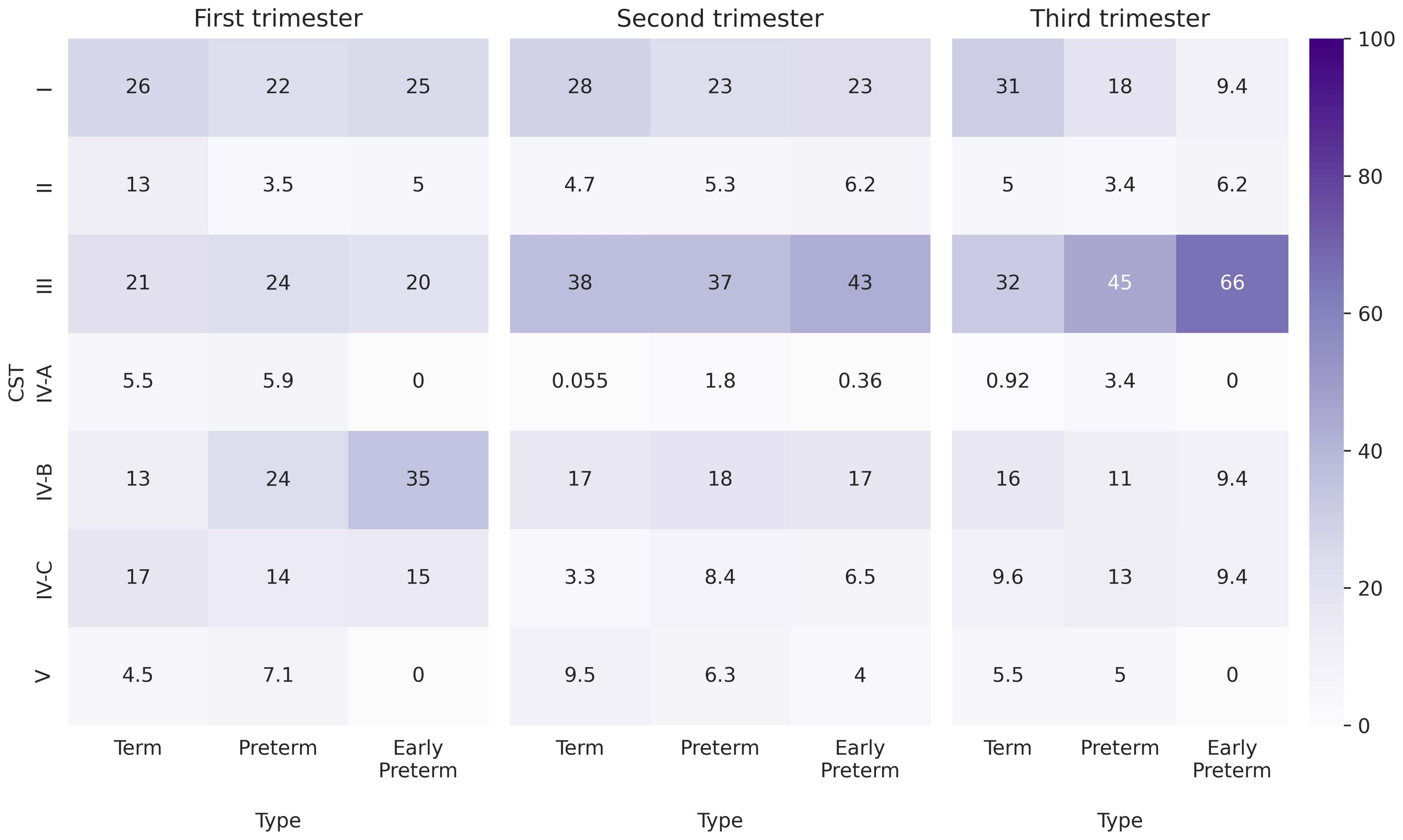

**Figure S4**: Subchallenge 1 (PTB) Boostrapped under- and over-sampled and inverted AUROC and AUPRC results by team.

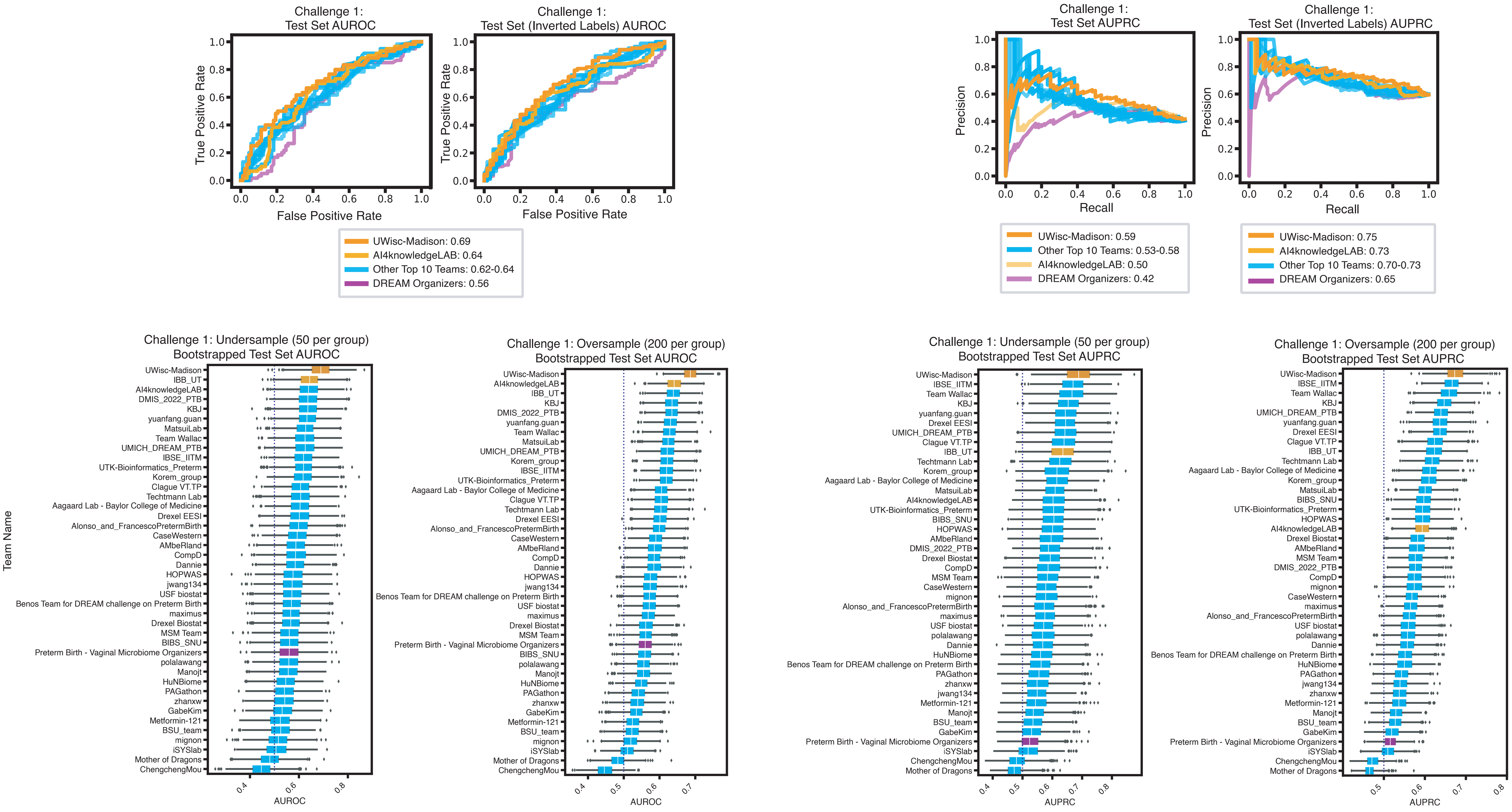

**Figure S5**: Subchallenge 2 (Early PTB) Boostrapped under- and over-sampled and inverted AUROC and AUPRC results by team.

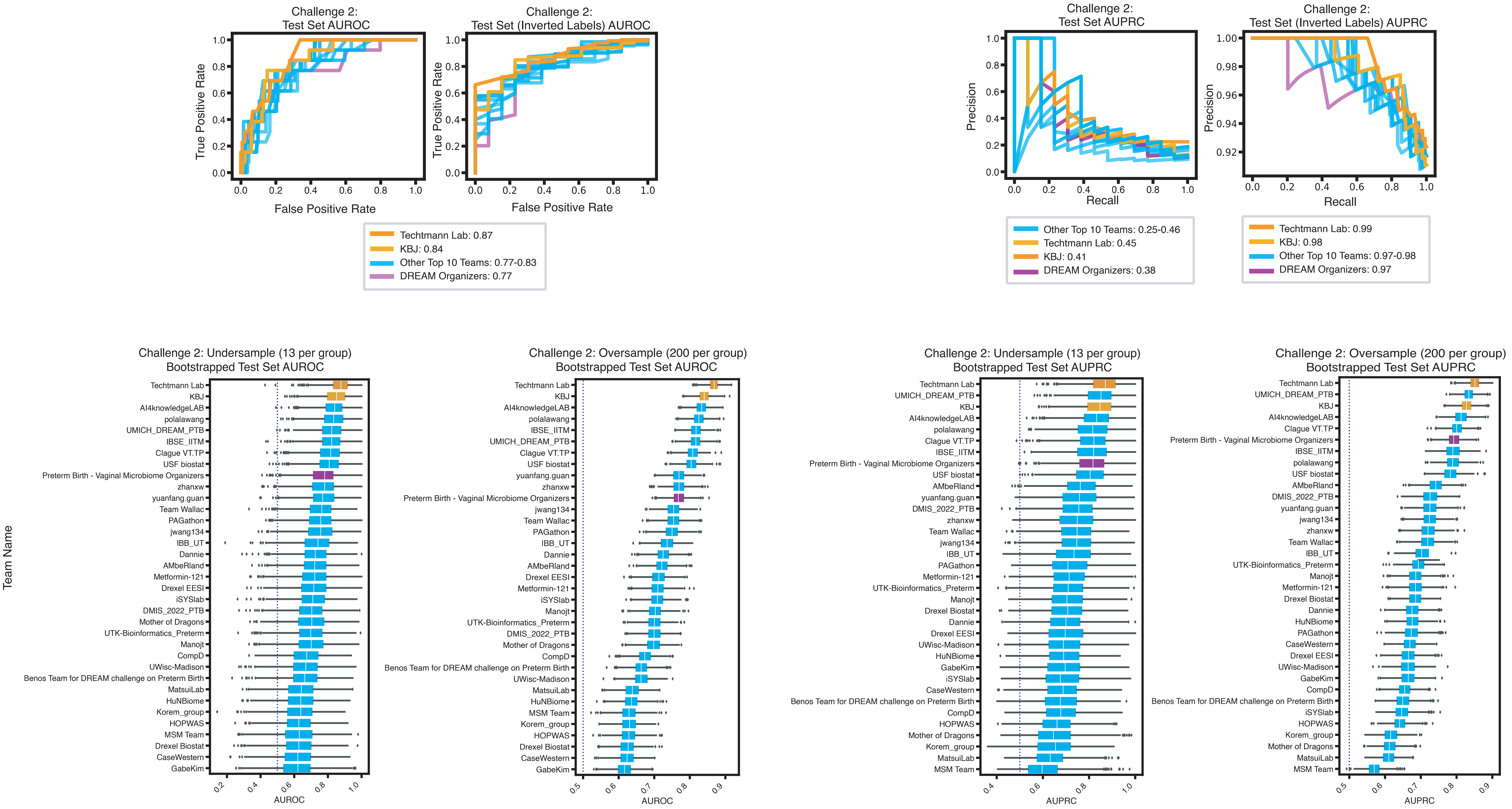

**Figure S6: Team UWisc Subchallenge 1 Overview**

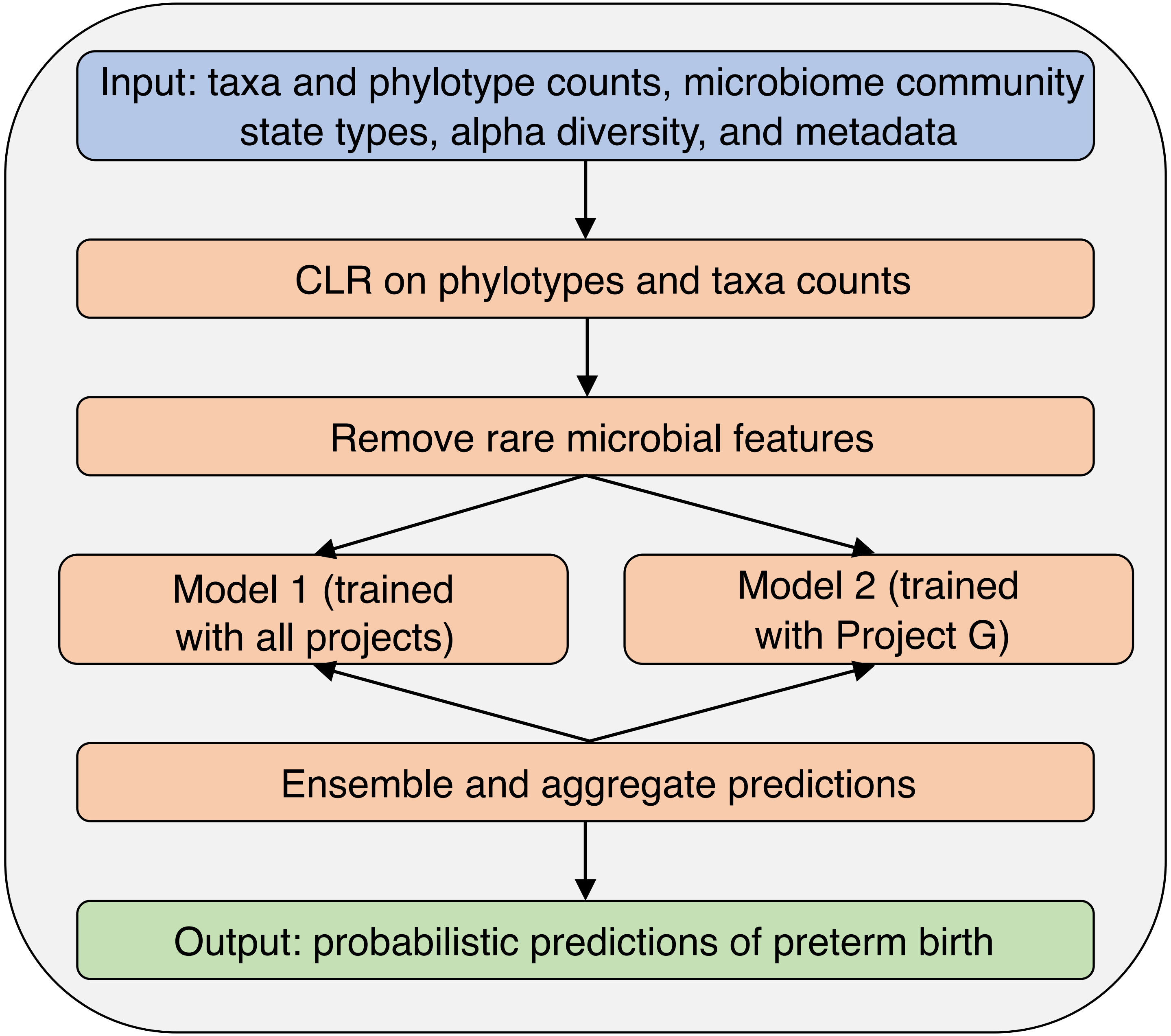

**Figure S7: AIKnowledgeLab Subchallenge 1 Overview**

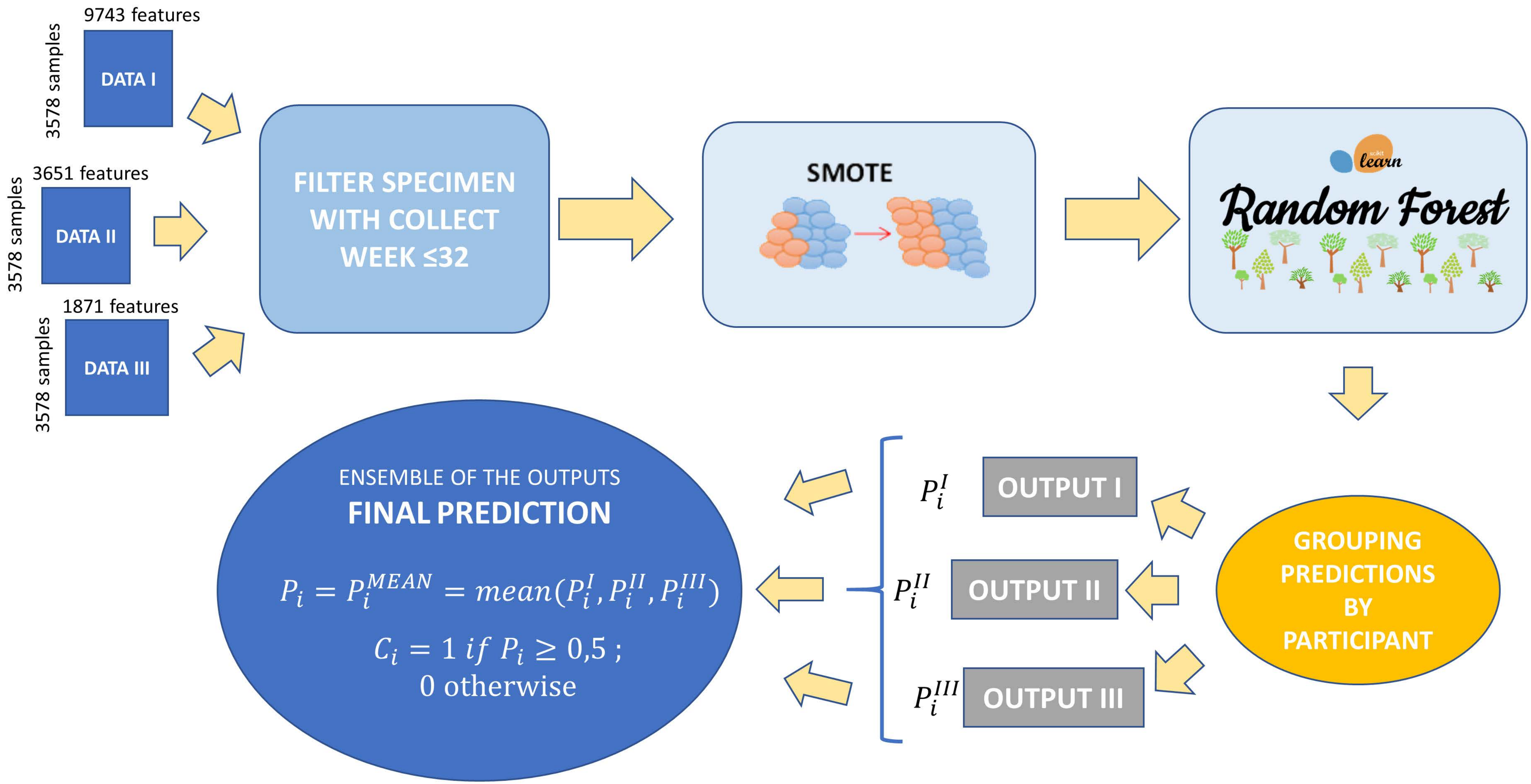

### Figure S8: KBJ Subchallenge 2 Overview

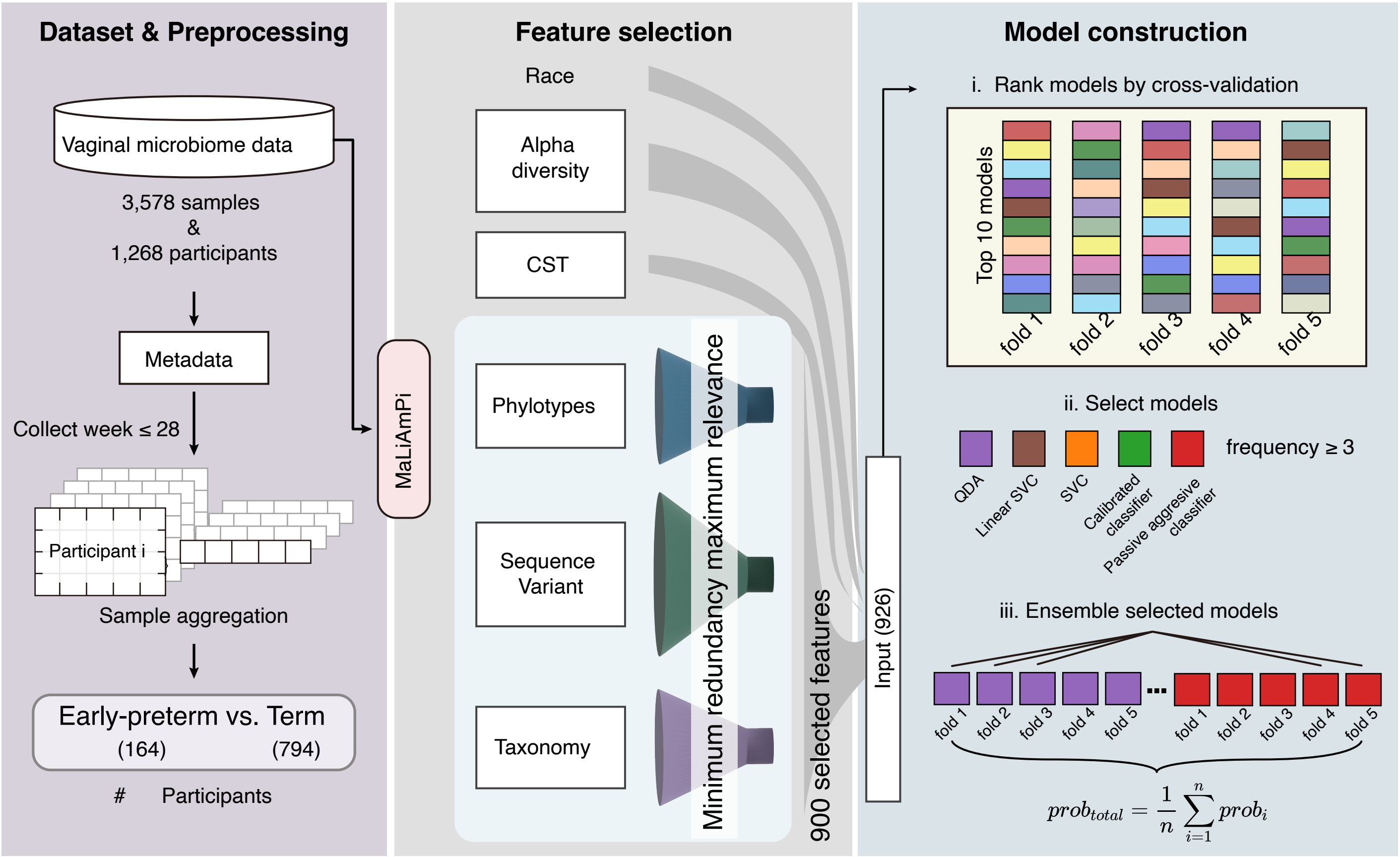

### Figure S9: Subchallenge 1 Feature Permutation Results

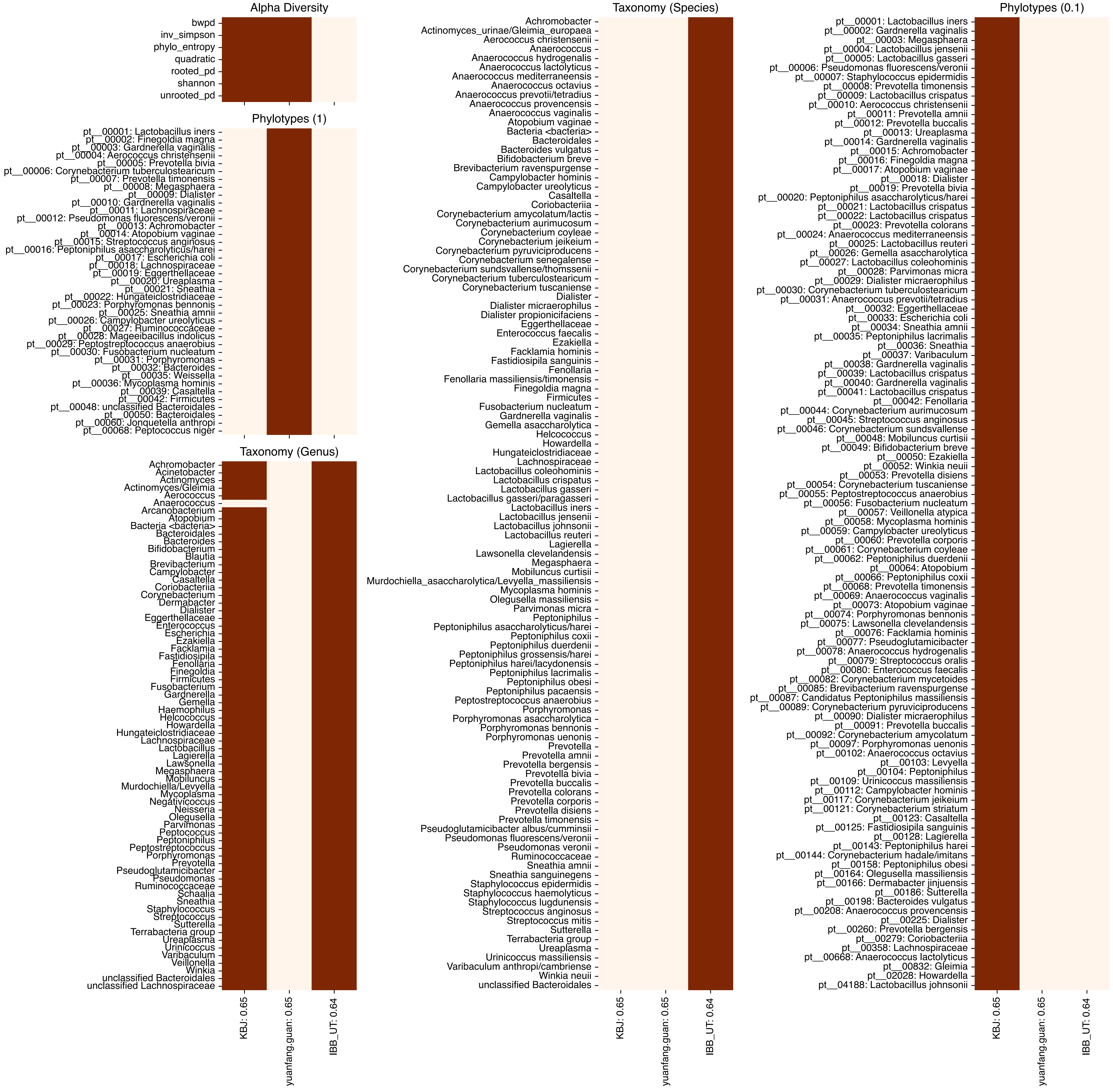

**Figure S10: Subchallenge 2 Feature Permutation Results**

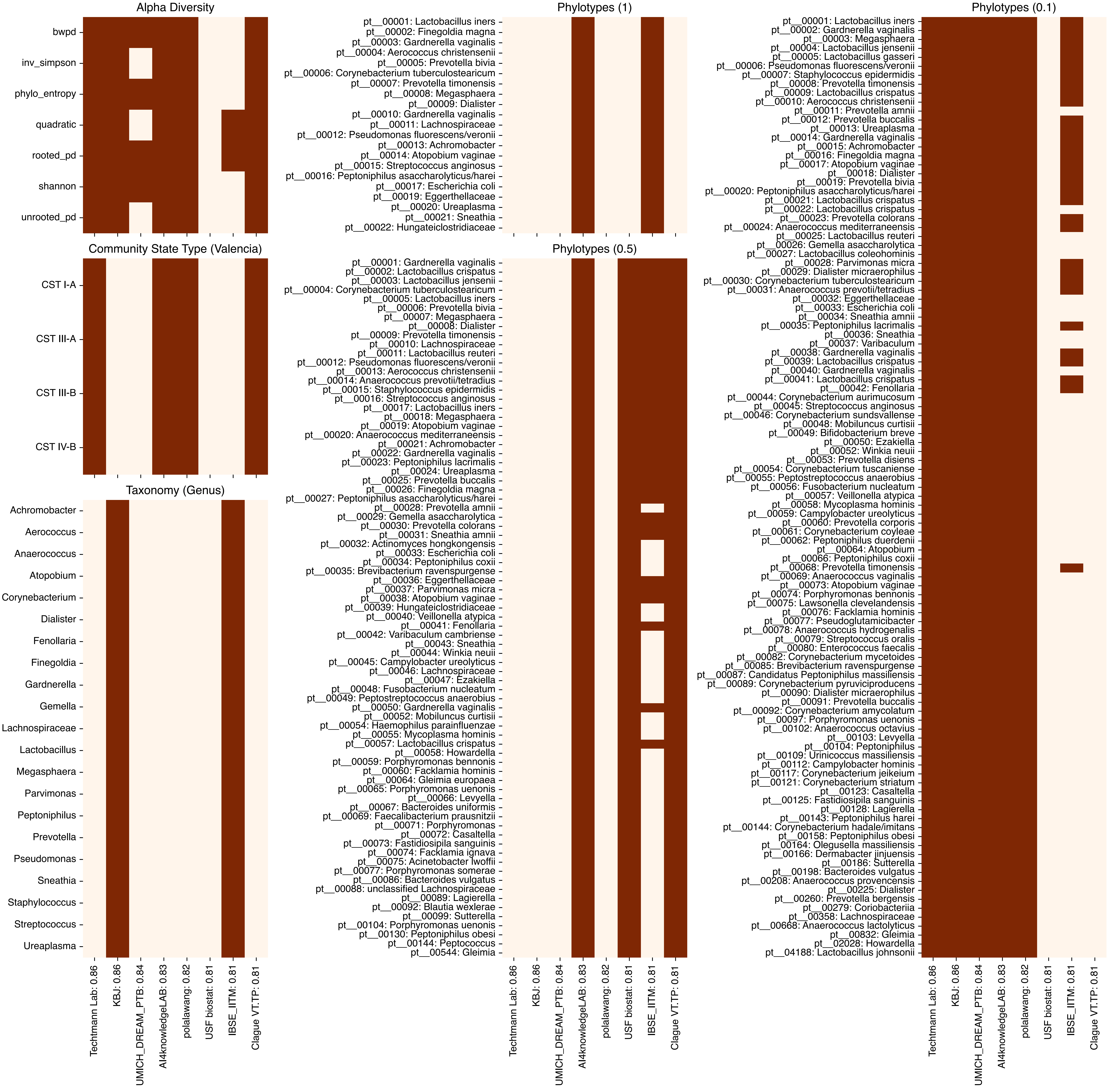
